# Leveraging Pretrained Models for Multimodal Medical Image Interpretation: An Exhaustive Experimental Analysis

**DOI:** 10.1101/2024.08.09.24311762

**Authors:** Temitayo Matthew Fagbola, Igwebuike Success

## Abstract

Artificial intelligence (AI) in radiology, particularly pretrained machine learning models, holds promise for overcoming image interpretation complexities and improving diagnostic accuracy. Although extensive research highlights their potential, challenges remain in adapting these models for generalizability across diverse medical image modalities, such as Magnetic Resonance Imaging (MRI), Computed Tomography (CT), and X-rays. Most importantly, limited generalizability across image modalities hinders their real-world application in diverse medical settings. This study addresses this gap by investigating the effectiveness of pretrained models in interpreting diverse medical images. We evaluated ten state-of-the-art convolutional neural network (CNN) models, including ConvNeXtBase, EfficientNetB7, VGG architectures (VGG16, VGG19), and InceptionResNetV2, for their ability to classify multimodal medical images from brain MRI, kidney CT, and chest X-ray (CXR) scans. Our evaluation reveals VGG16’s superior generalizability across diverse modalities, achieving accuracies of 96% for brain MRI, 100% for kidney CT, and 95% for CXR. Conversely, EfficientNetB7 excelled in brain MRI with 96% accuracy but showed limited generalizability to kidney CT (56% accuracy) and CXR (33% accuracy), suggesting its potential specialization for MRI tasks. Future research should enhance the generalizability of pretrained models across diverse medical image modalities. This includes exploring hybrid models, advanced training techniques, and utilizing larger, more diverse datasets. Integrating multimodal information, such as combining imaging data with patient history, can further improve diagnostic accuracy. These efforts are crucial for deploying robust AI systems in real-world medical settings, ultimately improving patient outcomes.

## 1. Introduction

In modern healthcare, medical imaging is an indispensable cornerstone, providing profound insights into the intricate structures and potential anomalies within the human body. The successful interpretation of medical images across varied modalities—such as CT, MRI, and X-rays—traditionally falls under the expertise of experienced radiologists. However, this interpretative process is multifaceted, marked by complexities inherent in analyzing medical images, from detecting subtle cues to providing comprehensive clinical evaluations due to increased image analysis demands (Pesapane et al., 2018; Balabanova et al., 2005). Each imaging modality has unique strengths and limitations, adding intricacies to its analysis. For instance, while CT scans offer detailed information, X-rays, especially chest X-rays (CXRs), are more cost-effective, expose patients to less radiation, and are more accessible (Power et al., 2016). These factors make CXRs particularly practical in resource-limited settings. Additionally, MRI can sometimes be a more suitable alternative for specific medical images, showcasing its potential to complement or substitute other modalities. In some cases, combining multiple imaging modalities improves accuracy and outcomes, recognizing the limitations of relying solely on a single modality (Puderbach et al., 2007; Abhisheka et al., 2023). This convergence exemplifies the evolving complexity of medical image interpretation.

Moreover, the increasing demand for radiological investigations is juxtaposed against a diminishing number of radiologists, creating an imbalance that underscores the urgent need for innovative solutions to augment diagnostic capacities. In this context, the evolving role of artificial intelligence (AI) in healthcare has emerged as a beacon of promise (Biswas et al., 2019). AI applications, from image analysis to diagnostic assistance and predictive modelling, hold the potential to significantly enhance outcomes across diverse domains, including patient care (Ben-Israel et al., 2020). Particularly noteworthy is the concept of pre-trained models within AI. Pre-trained models represent a paradigm shift in machine learning, wherein models are initially trained on large and diverse datasets to learn generalized features. This learned knowledge is subsequently repurposed, fine-tuned, and adapted to specific tasks or domains.

In medical image analysis, leveraging these pre-trained models offers a spectrum of potential benefits (Litjens et al., 2017). These models have showcased remarkable performance across various medical image modalities, excelling in both binary and multiclass classification tasks. Models like ResNet, VGG, and DenseNet, initially trained on large-scale natural image datasets, have demonstrated transferability and robustness in classifying medical images. Their ability to extract hierarchical features underscores their adaptability and effectiveness. Notably, these models have achieved high accuracies, sensitivities, and specificities in diagnosing conditions like tumors, fractures, and abnormalities, thus proving their efficacy in diverse medical imaging scenarios. However, continual validation across varied datasets and clinical scenarios is pivotal to ensure consistent and reliable performance across different modalities and classification tasks. The widespread use and effectiveness of pre-trained models in analyzing medical images have sparked interest in understanding how well these models can adapt to different scenarios. In response to this interest, we have developed an evaluation framework designed to assess the performance and adaptability of pre-trained models—including some proposed in previous research studies— particularly in the context of interpreting medical images.

The contribution of this study is as follows:

i. We evaluate the diagnostic accuracy of ten pretrained models across diverse medical imaging modalities, using three datasets containing MRI scans, CT scans, and X-rays. This assessment provides insights into the models’ ability to generalize and adapt to different types of medical images.
ii. We explore two distinct classification scenarios to enhance the effectiveness of multi-label diagnosis in medical images. First, we implement a four-category classification approach for two datasets, each corresponding to a different modality. Second, we employ a three-category classification approach, resulting in more accurate and efficient diagnoses in medical settings.
iii. We evaluate the robustness and reliability of the pretrained models, ensuring consistent performance across diverse datasets and imaging modalities. This consistency is crucial for dependable clinical decision-making.
iv. We identify and address biases present in pretrained models, ensuring unbiased and equitable diagnoses across multiple imaging types. This contributes to fairness in medical imaging.

The rest of this paper is structurally organized as follows: Section 2 presents the relevant background and related works; Section 3 elaborates on the materials and methods employed, encompassing aspects such as data acquisition, preprocessing, algorithm selection, and evaluation metrics. Section 4 presents the outcomes of the study, unveiling the results obtained from the proposed framework. Following this, Section 5 engages in discussion, analysing and contextualizing the findings within the broader scope of research. Finally, Section 6 offers a conclusion, summarizing the key insights gleaned from the study.

## 2. Related Works

Over the years, researchers have shown an increased interest in leveraging machine learning algorithms for medical imaging analysis, particularly in diagnosing and assessing treatment responses. In this context, a notable study Abirami A. (2023) introduces EfficientNetB7 as a pre-trained deep learning model aimed at enhancing accuracy while reducing complexity. Utilizing a dataset comprising 3674 MRI images, the research focuses on evaluating the model’s efficacy in classifying tumors as glioma, meningioma, pituitary tumors, or non-tumorous. Leveraging transfer learning, the strategy implemented with EfficientNetB7 yielded a remarkable 98.4% accuracy. However, it’s important to note that this high accuracy was achieved without evaluation against other datasets or modalities, raising questions about the model’s generalizability beyond the specific dataset used in the study. The study by Abdelaziz Ismael et al. (2020a) introduces an improved deep learning model designed for brain tumor classification using MRI images. The model was tested on a benchmarked dataset comprising 3064 MRI images across three tumor types. It achieved a remarkable accuracy of 99%, surpassing previous benchmarks on the same dataset.

Ramadhan & Baykara (2022) introduced an updated VGG16-CNN model of reduced parameters from approximately 138 million parameters to around 40 million parameters for multiple classifications of COVID-19, two of which classification comprised of three classes: COVID-19, normal, and pneumonia, and binary classification of COVID-19 and normal class. The research utilized three datasets: Db1 containing 21,165 images, Db2 with 5,226 images, and Db3 comprising 6,432 images. Results were promising, with high accuracy achieved across the datasets. Specifically, the model attained 99% accuracy for triple classification and 100% for binary classification on Db3, 98% and 99% on Db2, and 96% and 92% on Db1 for triple and binary classifications, respectively. In another study, a novel model named DarkCovidNet Ozturk et al. (2020) is introduced for the automatic detection of COVID-19 using raw chest X-ray images. The model is specifically designed for accurate diagnostics in both binary classification (COVID vs. No-Findings) and multi-class classification (COVID vs. No-Findings vs. Pneumonia) scenarios. The development phase utilized a dataset comprising 1125 images, categorized into 125 COVID-19 positive, 500 Pneumonia, and 500 No-Findings cases.

Remarkably, the model achieved an accuracy of 98.08% for binary classification and 87.02% for multi-class classification. The DarkNet model employed in the study served as a classifier for the you only look once (YOLO) real-time object detection system. Limitations of these studies, such as the relatively small dataset and the need to expand their research by exploring larger datasets and other medical image types, are acknowledged, they also address the imperative of evaluating competitive models across varied medical images, highlighting their focus on ensuring the model’s robustness and accuracy—a pivotal aspect that aligns with the overarching aim and contribution of this study. Leveraging pretrained models may herald advancements in diagnostic accuracy and efficiency, hinting at a transformative paradigm for medical imaging interpretation and augmenting clinical decision-making processes. Table 1 showcases the instrumental role of pretrained models in advancing technological capabilities, as demonstrated by several related studies across diverse domains.

**Table 1.** Related Works in the application of pretrained model for medical image classification.

| Methodology | Number of Dataset & Modality | Classification | Model |
| --- | --- | --- | --- |
| Narin et al. (2021) | CXR datasets (3) | Multi – 3 classes | ResNet50 |
| Kassania et al. (2021) | CXR & CT images | Binary | DenseNet121 + Bagging Tree classifier |
| Monowar et al. (2020) | CXR | Multi – 3 classes | Xception |
| Xue et al. (2023) | CXR & CT images | Multi – 3 classes | Ensemble: VGG16, DenseNet, ResNet50, ResNet102 |
| Alshmrani et al. (2023) | CXR | Multi – 6 classes | VGG19 + CNN |
| Alshmrani et al. (2023) | CXR | Binary & Multi – 3 classes | Modified ResNet50 |
| Raza et al. (2023) | CT images | Multi – 3 classes | Lung-EffNet |
| Humayun et al. (2022) | CT images | Multi – 3 classes | VGG16, VGG19, & Xception |
| Zhou et al. (2023) | CT images | Multi – 3 classes | Ensemble |
| Ibrahim et al. (2023) | CT images (2) | Binary | COV-CAF |
| Jangam et al. (2022) | CXR (2) & CT images (3) | Binary | Stacked Ensemble |
| Abdelaziz Ismael et al. (2020) | MRI | Multi – 3 classes | Residual Networks |
| Nayak et al. (2020) | MRI datasets (2) | Multi – 5 classes | Custom CNN |

## 3. Materials and Method

A systematic selection process identified a cohort of leading pretrained models alongside benchmarked architectures for comparative evaluation. These models encompass a spectrum of ten Convolutional Neural Network (CNN) architectures, including DenseNet201, NasNetMobile, NasNetLarge, Xception, ResNet50, EfficientNetB7, VGG16, VGG19, InceptionResNetV2, ConvNeXtBase and four benchmarked models namely; VGG19+CNN, Modified ResNet50, Custom CNN and Ensemble stacked models. The adaptability of these fourteen models across diverse medical imaging modalities was rigorously assessed for their capacity to interpret CT scans, MRIs, and X-rays effectively. The evaluation framework employed a diverse set of metrics, gauging model performance across specific medical imaging tasks, emphasizing diagnostic accuracy, precision, and sensitivity. In Figure 1, the workflow for the experimental analyses is presented.

**Figure 1:**
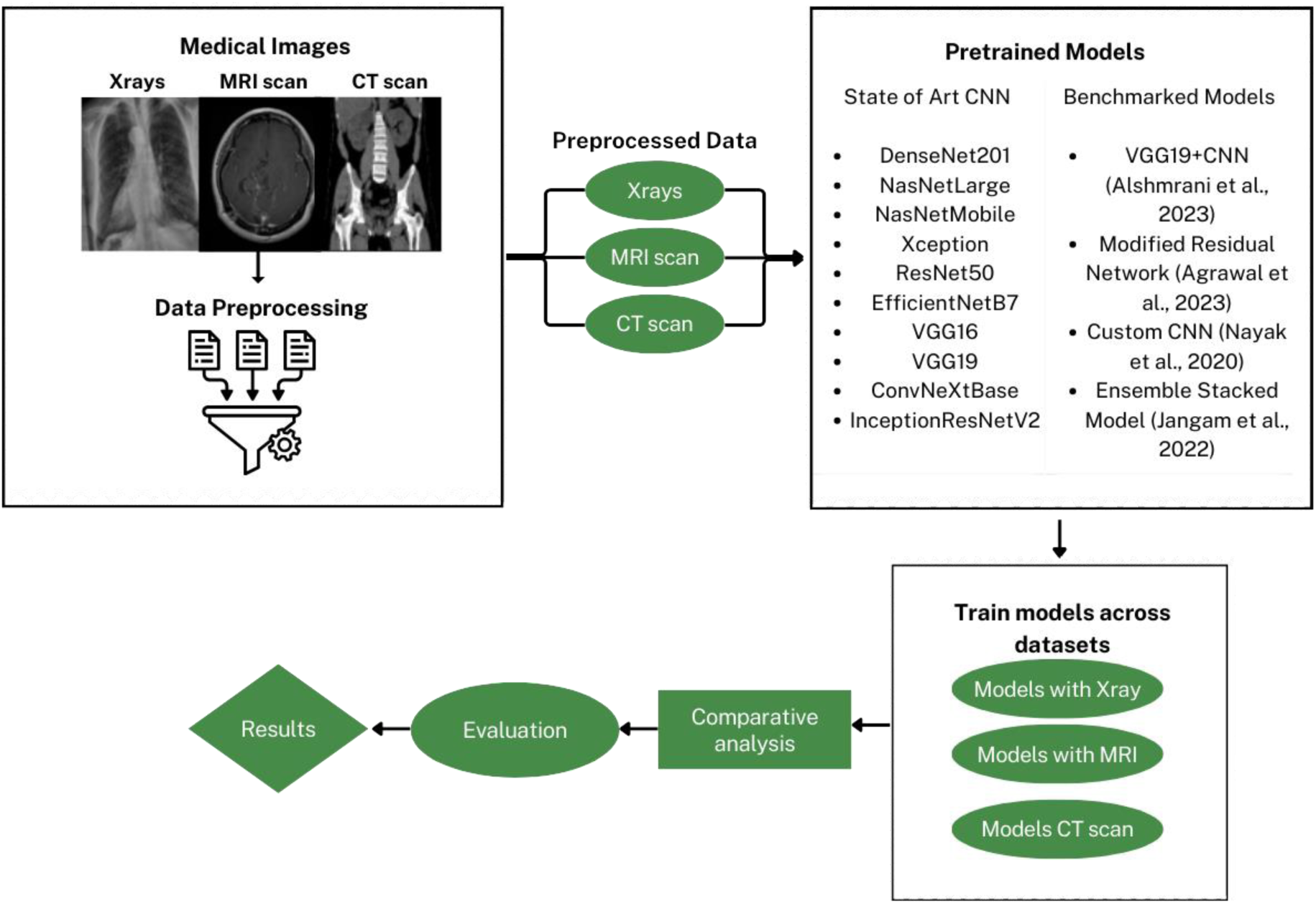
Overall Architecture of the Proposed Framework.

### 3.1 Datasets

1. The Brain MRI dataset (Nickparvar, 2021), used for this study encompasses 7023 images sourced from publicly available datasets like figshare, Br35H, and SARTAJ. These images are categorized into glioma, meningioma, no tumor, and pituitary classes. The dataset was split into train and test set with the 80:20 ratio and a separate set of images was used for validation. Various researchers have utilized this dataset for medical image classification tasks. For example, Özkaraca et al. (2023) developed a new deep learning model for MR image classification, leveraging strengths of DenseNet, VGG16, and basic CNNs. Similarly, Islam *et al*. (2023) explores deep transfer learning architectures for brain tumour diagnosis using MRI.
2. The <u>Kidney</u> CT Scan dataset includes 12,446 images representing tumor, cyst, normal, and stone classes. The dataset underwent preprocessing that involved identifying and removing duplicates using a robust hashing method, resulting in a refined set of 11,929 images. These images were split into ratio of 70:20:10 for training, testing, and validation. The original 512×512×3 pixel images were resized to 224×224×3 pixels to balance model accuracy and computational complexity for efficient model training.
3. The <u>Chest</u> X-ray Dataset for analysis consisted of samples from nine different sources, including augmented images to compensate for the limited availability of a single extensive dataset. It comprises three primary classes: covid, pneumonia, and normal cases, totaling 6,939 samples. This diverse dataset enabled a comprehensive analysis of chest X-ray instances. The dataset underwent preprocessing, transitioning into a structured dataframe. It was then divided into 80% for the training set and 20% for the testing set. The images were standardized to a 224×224×3 dimension using rescaling and resizing techniques. This resizing aimed to enhance the model’s focus by reducing irrelevant information assimilation from larger images, promoting a more pertinent learning process. Table 2 illustrates the breakdown of image proportions utilized for model evaluation.

**Figure 2:**
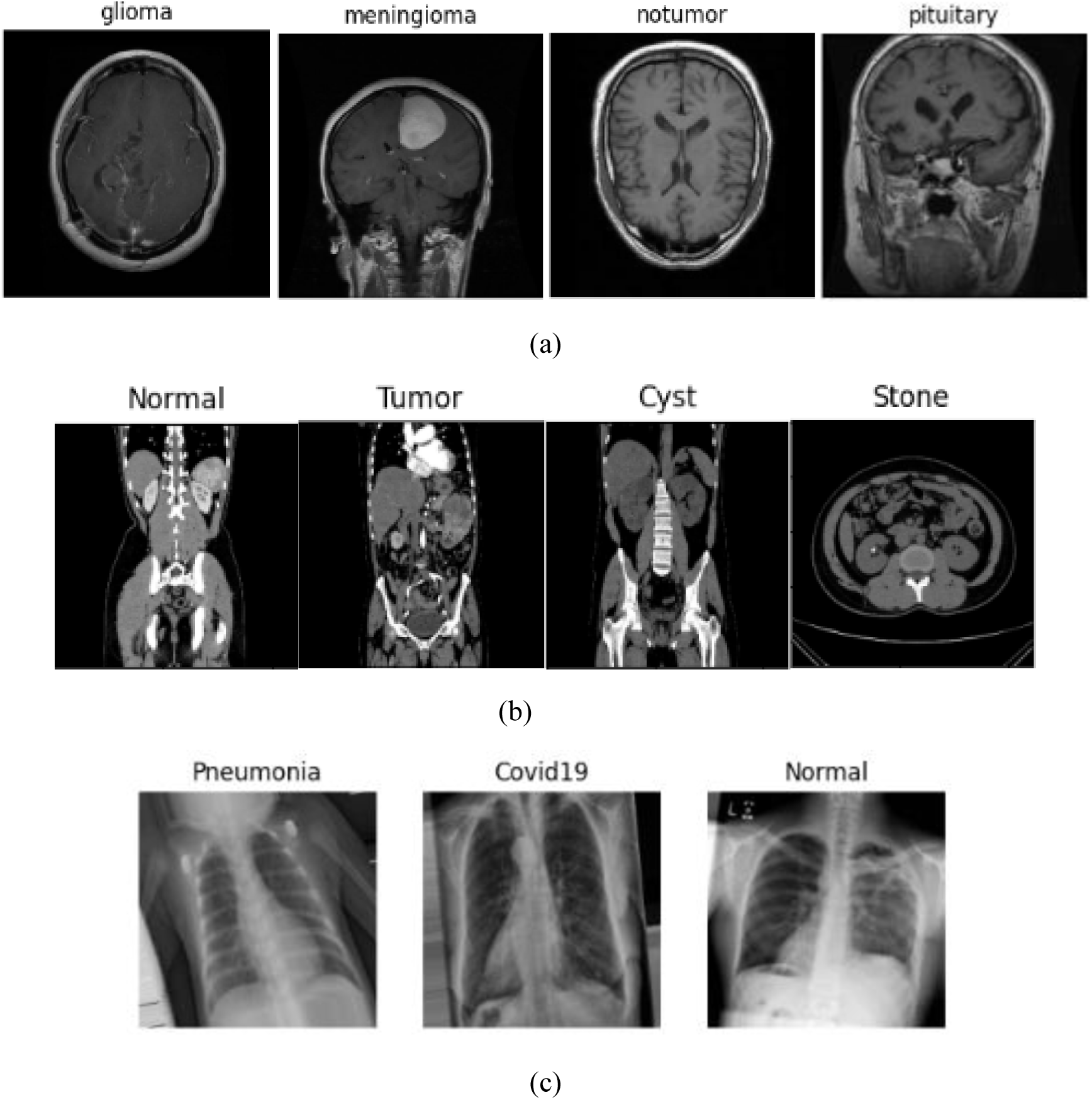
Dataset Snippet Arranged by Modalities a, b, and c –Brain MRI, Kidney CT, and CXR Images respectively.

### 2.2 Algorithm Background

1. **DenseNet201:** DenseNet-201, (Huang et al., 2022), a specific variant of the DenseNet architecture. DenseNet-201 is renowned for its unique structure, characterized by dense connectivity patterns among layers. In a DenseNet, each layer receives inputs from all preceding layers, promoting extensive feature reuse and propagation. This dense connectivity enhances gradient flow during training, mitigating issues like vanishing gradients and enabling more efficient learning. Due to its densely connected layers, DenseNet-201 can effectively capture intricate dependencies between features within the data.
2. **NasNetLarge and NasNetMobile:** NasNet (Neural Architecture Search Network), models are designed using neural architecture search techniques, resulting in architectures optimized for performance and efficiency. the creation of a novel search space, referred to as the “NASNet search space,” refers to a predefined set of possible neural network architectures or architectural components that are explored during the process of neural architecture search (NAS), (Zoph et al., 2018).
3. **Xception:** Xception derived from the combination of “Extreme Inception” signifies an extreme iteration of the Inception architecture, a well-known convolutional neural network design employing “Inception modules” for feature extraction. In Xception, these Inception modules are replaced with “depthwise separable convolutions,” a variant of convolutions that separates spatial and channel-wise information processing. This alteration in convolutional operation distinguishes Xception from Inception. The Xception architecture is characterized by its 36 convolutional layers, serving as the foundational feature extraction component of the network (Chollet, 2017a).
4. **Vgg16 & Vgg19:** VGG (Visual Geometry Group) networks are characterized by their simple and uniform architecture, comprising multiple convolutional layers followed by max-pooling and fully connected layers. The authors (Simonyan & Zisserman, 2015) evaluated convolutional neural networks (CNNs) of increasing depth, up to 16–19 weight layers, using a specific architecture with small (3×3) convolution filters, which positively impacts classification accuracy. VGG16 and VGG19 differ in depth, with VGG19 having more layers. These models are chosen for their simplicity, ease of interpretation, and strong performance.
5. **EfficientNetB7:** EfficientNet employs a compound scaling method to balance model depth, width, and resolution, resulting in highly efficient yet powerful architectures. EfficientNetB7 is the largest variant in the EfficientNet family, offering superior performance on various computer vision tasks while maintaining high efficiency. Thanks to this compound scaling method, a mobile-sized EfficientNet model can be scaled up very effectively, achieving state-of-the-art accuracy with significantly fewer parameters (Tan & Le, 2019).
6. **InceptionResNetV2:** InceptionResNetV2, a combination of Inception architectures with residual connections, leverages the benefits of both approaches. It is a costlier hybrid Inception version but despite the increased computational cost, the model demonstrates better accuracy or effectiveness in recognizing and classifying objects in images compared to previous versions or other architectures (Szegedy et al., 2017).
7. **Resnet50:** ResNet (Residual Network) introduced residual connections to address the vanishing gradient problem in deep neural networks. The ResNet architecture consists of multiple “blocks” of layers, each containing two convolutional layers followed by batch normalization and a ReLU activation function. ResNet50 is a specific variant with 50 layers, each 2-layer block in the 34-layer net was replaced with this 3-layer bottleneck block, resulting in a 50-layer ResNet, striking a balance between depth and computational complexity (He et al., 2015).
8. **ConvNeXtBase:** ConvNeXtBase employs grouped convolutions to enhance feature extraction while reducing computational cost. ConvNeXts were constructed using standard ConvNet modules, and they perform well in comparison to Transformers across various metrics such as accuracy, scalability, and robustness on major benchmarks. Despite being based on traditional ConvNet modules, ConvNeXts are competitive with Transformers, which represent another type of neural network architecture (Liu et al., 2022).

**Figure 3:**
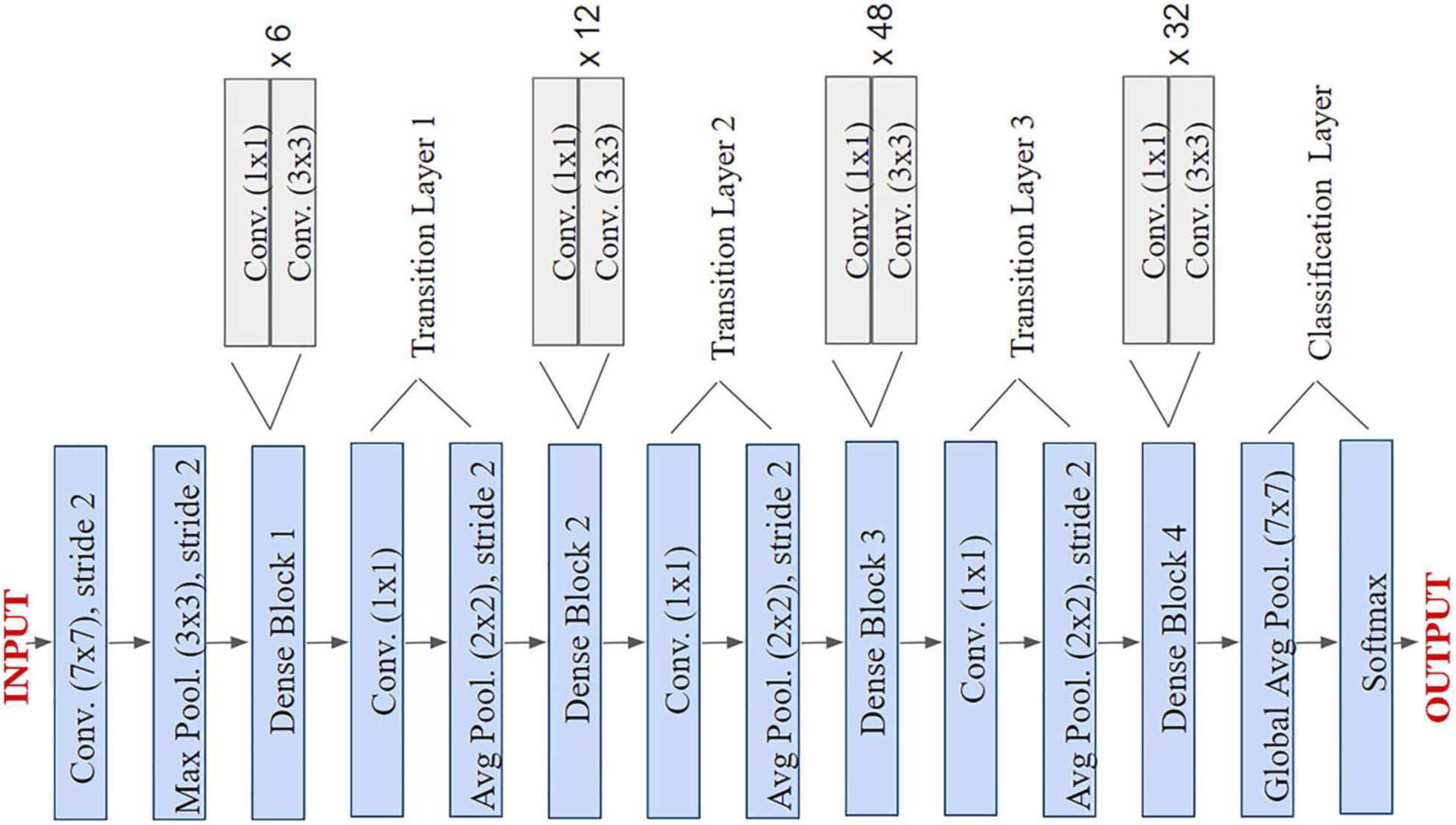
The Architecture of DenseNet201 (Chahar et al., 2020).

**Figure 4:**
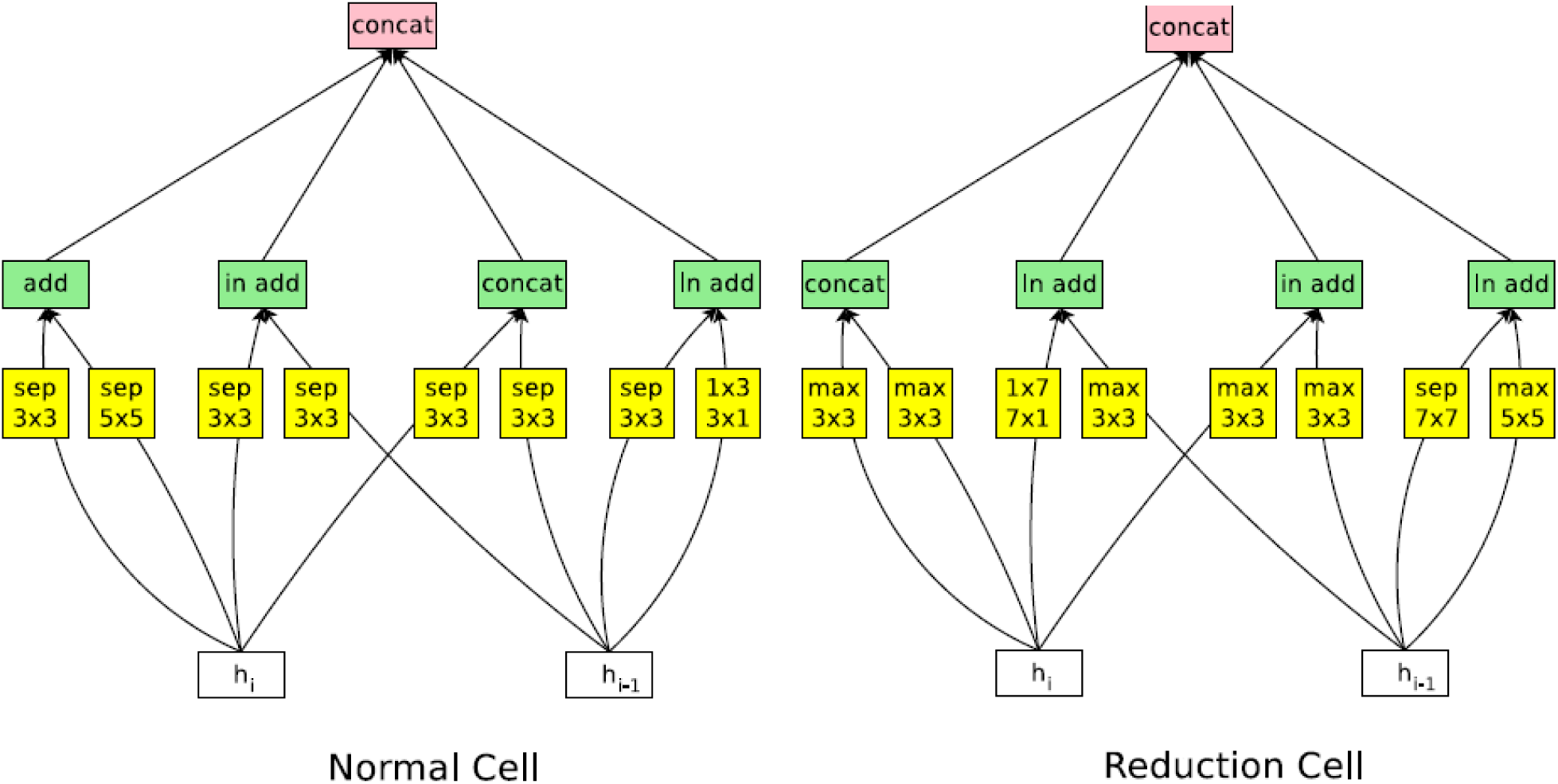
The Architecture of NasNet (Tsang, 2021).

**Figure 5:**
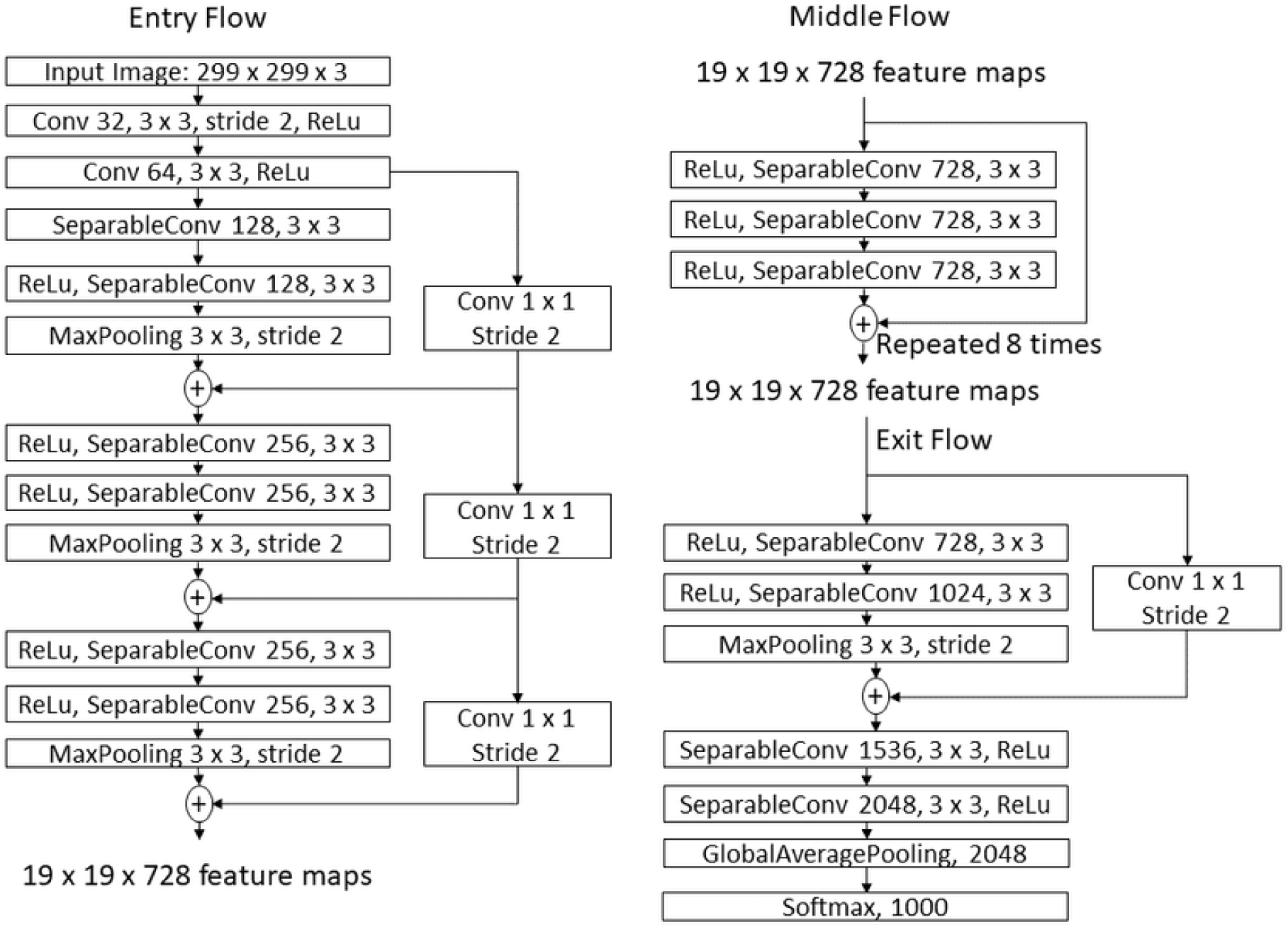
The Architecture of Xception (Chollet, 2017b).

**Figure 6:**
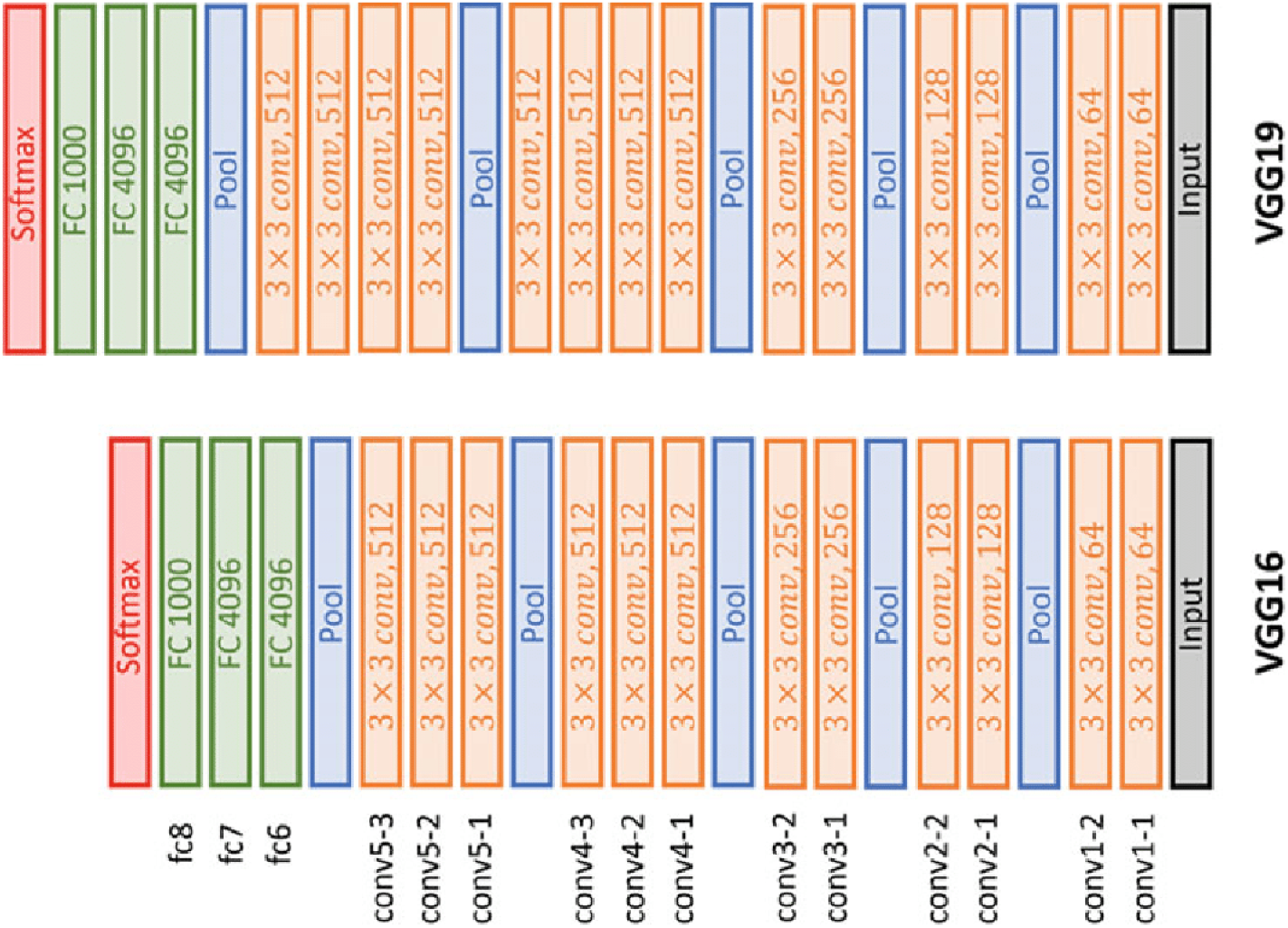
The Architecture of VggNet (El-Rahiem & Hammad, 2021).

**Figure 7:**
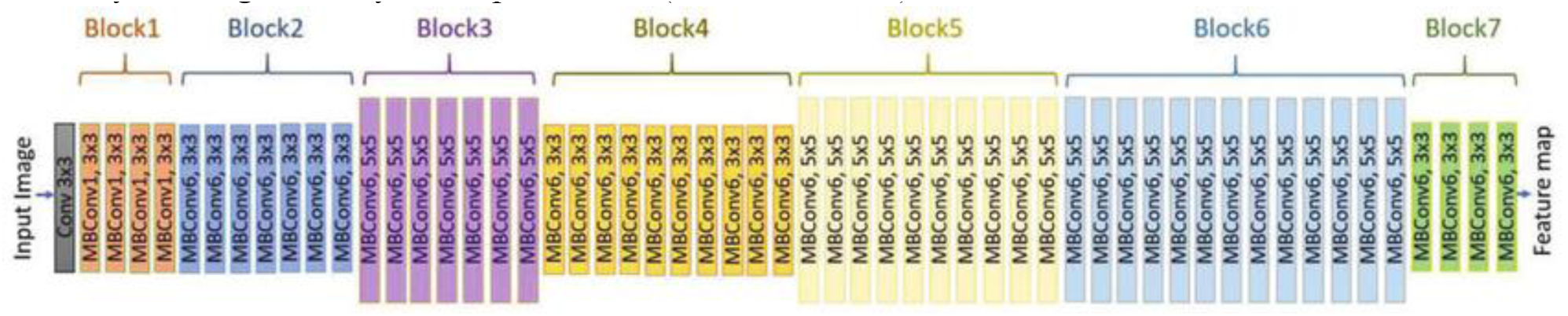
EfficientNetB7 Model Architecture (Tan & Le, 2019).

**Figure 8:**
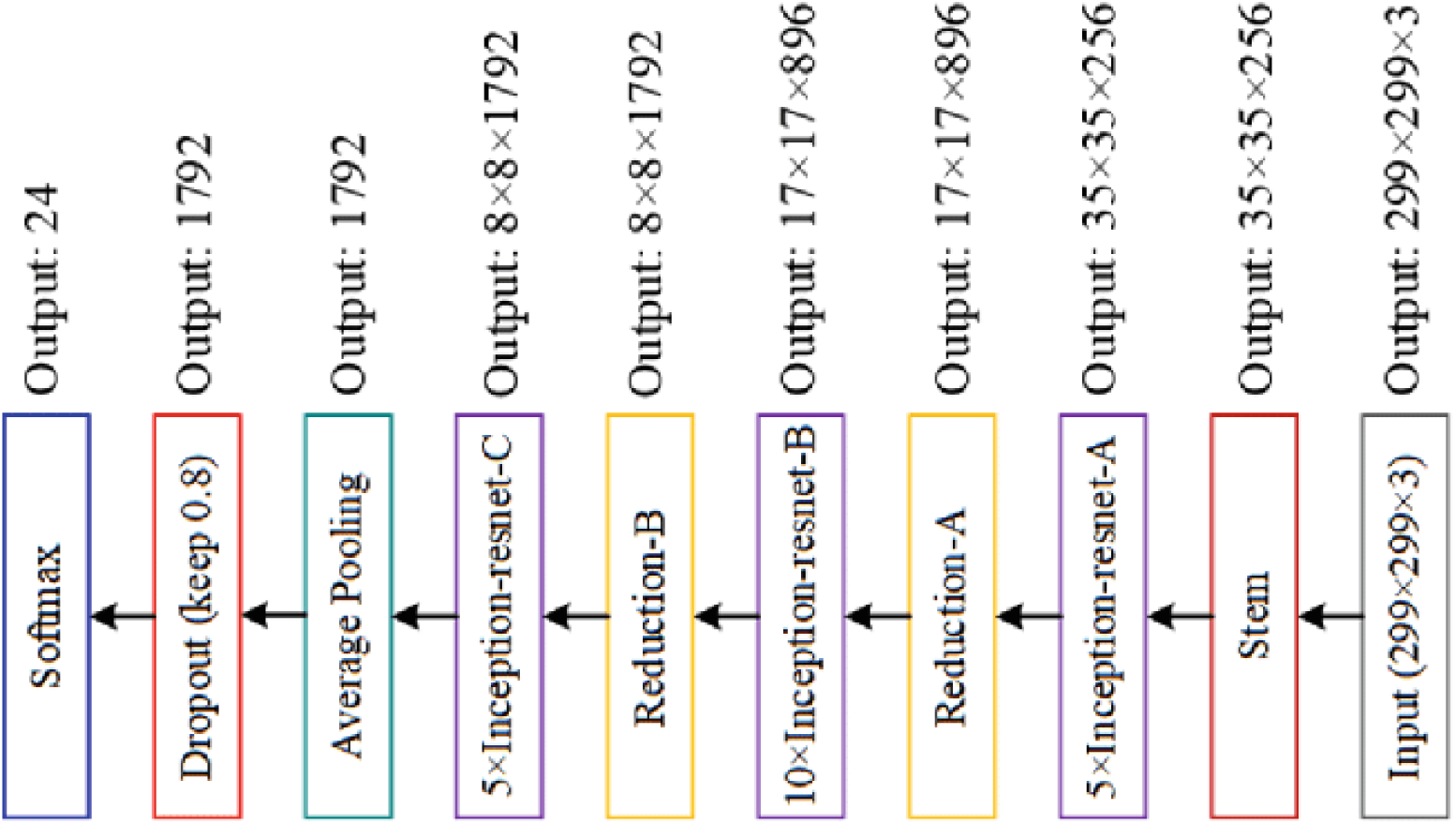
The Architecture of InceptionResNetV2 (Gao et al., 2020).

**Figure 9:**
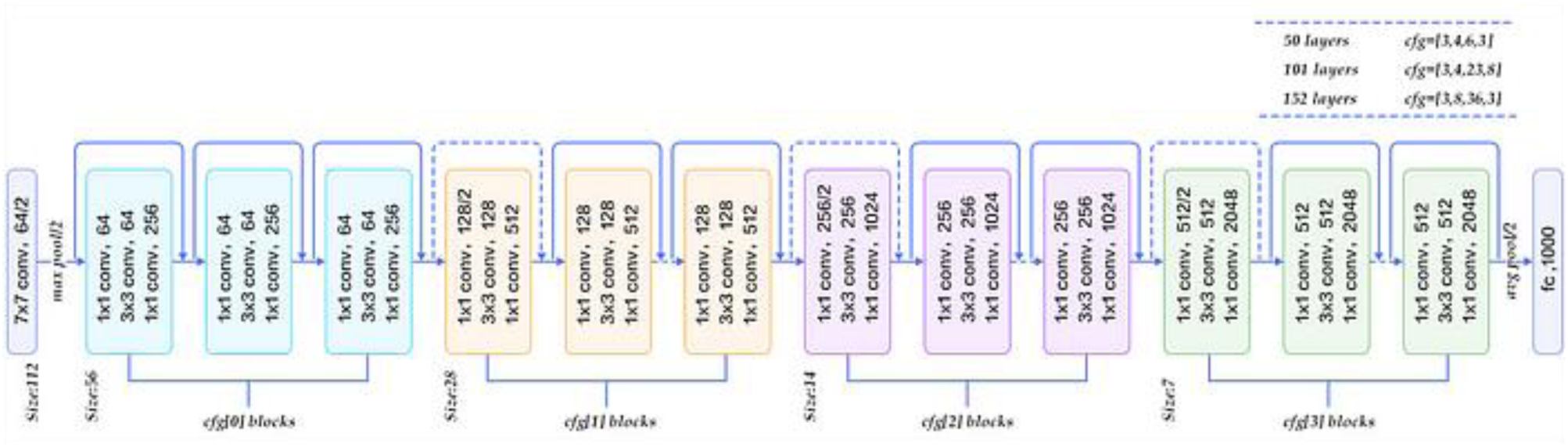
The Architecture of Resnet50 (Rastogi, 2022).

**Figure 10:**
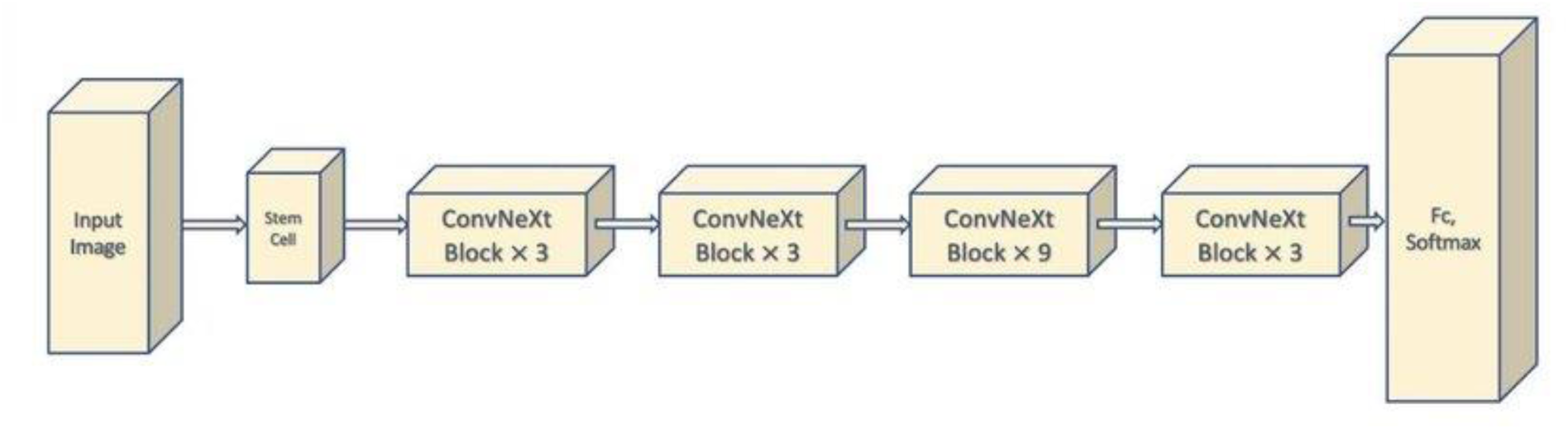
The CovNeXt Architecture (Yengec-Tasdemir et al., 2022).

### 2.3 Experiments

Computational capabilities –the experiment was carried out by utilizing Google Colab’s V100 GPU configuration, boasting 40GB of RAM. Additionally, the Dell machine used is equipped with a Core i9 – 12900 CPU with 32GB of RAM and powered by an NVIDIA GeForce RTX 3090 GPU.

**Table 2.**
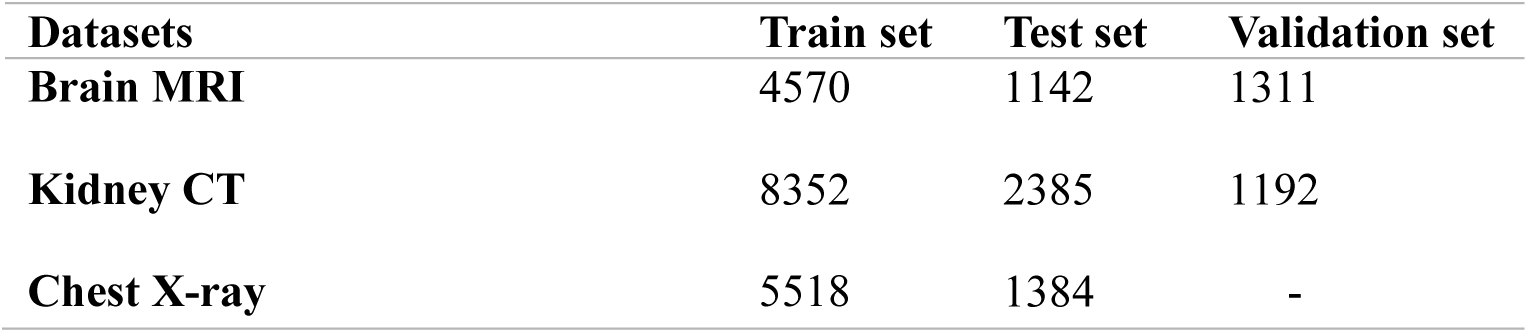
Proportional Distribution of Images for Model Evaluation.

For implementation, the state-of-the-art CNN models underwent initial training on the ImageNet dataset. Following image preprocessing, all layers of these pretrained models were frozen. To ensure uniformity and minimize bias among the models, no extra deep neural networks or dense layers were introduced, ensuring methodological consistency and fairness in model assessment, the input shape was meticulously set to match the size expected by most CNN pretrained models. For the optimization process, we opted for the Adam optimizer, a widely acclaimed choice known for its adaptive learning rate properties. This optimizer has been extensively used in various domains and is considered a standard choice in the machine learning community, also, adaptive nature of the learning rate in Adam ensures robust convergence and efficient training (Wang et al., 2022). In determining the appropriate duration for training, our selection of the number of epochs was guided by the need to witness the stabilization of model performance. By allowing sufficient epochs, we aimed to capture the convergence behaviour and ascertain the models’ adaptability. In the choice of batch size, we adhered to a parameter commonly employed in the literature. This careful selection ensures a balanced trade-off between stochastic updates and computational efficiency, aligning our approach with established practices for training deep neural networks. Table 3 provides an overview of the hyperparameters employed throughout the training process of the State of the Art pretrained models.

**Table 3.** Hyperparameters Used for State of Art CNN Models and Respective Values.

|  | Input Shape | Epochs | Batch Size | Optimizer | Learning Rate | Loss Function | Activation Function |
| --- | --- | --- | --- | --- | --- | --- | --- |
| **MRI** | 224x224 | 25 | 32 | Adam | 0.001 | Sparse categorical crossentropy | Softmax |
| **CT** | 224x224 | 25 | 32 | Adam | 0.001 | Sparse categorical crossentropy | Softmax |
| **CXR** | 224x224 | 25 | 32 | Adam | 0.001 | Categorical crossentropy | Softmax |

The hyperparameters of the benchmarked pretrained models from existing work were used for the training and evaluation process for the comparative analysis, as depicted in Table 4. This approach enables us to assess the performance of our proposed method against existing benchmarks under standardized conditions

**Table 4.** Hyperparameters Used for Benchmarked Models according to their specific architectures.

|  | **Input Shape** | **Epochs** | **Batch Size** | **Optimizer** | **Learning Rate** | **Loss Function** | **Activation Function** |
| --- | --- | --- | --- | --- | --- | --- | --- |
| **VGG19+CNN**<br>**Alshmrani et al. (2023)** | 224x224 | 5000 | 32 | Adam | 0.000009 | Sparse categorical crossentropy | Softmax |
| **MODIFIED RESNET**<br>**Agrawal et al. (2023)** | 224x224 | 25 | 32 | Adam | 0.001 | Sparse categorical crossentropy | Softmax |
| **CUSTOM CNN**<br>**Nayak et al. (2020)** | 224x224 | 10 | 25 | SGD | 0.0001 | Categorical crossentropy | Softmax |
| **ENSEMBLE STACKED MODEL**<br>**Jangam et al. (2022)** | 224x224 | 100 | 16 | Adam | 0.0001 | Categorical crossentropy | Softmax |

**Table 4.**
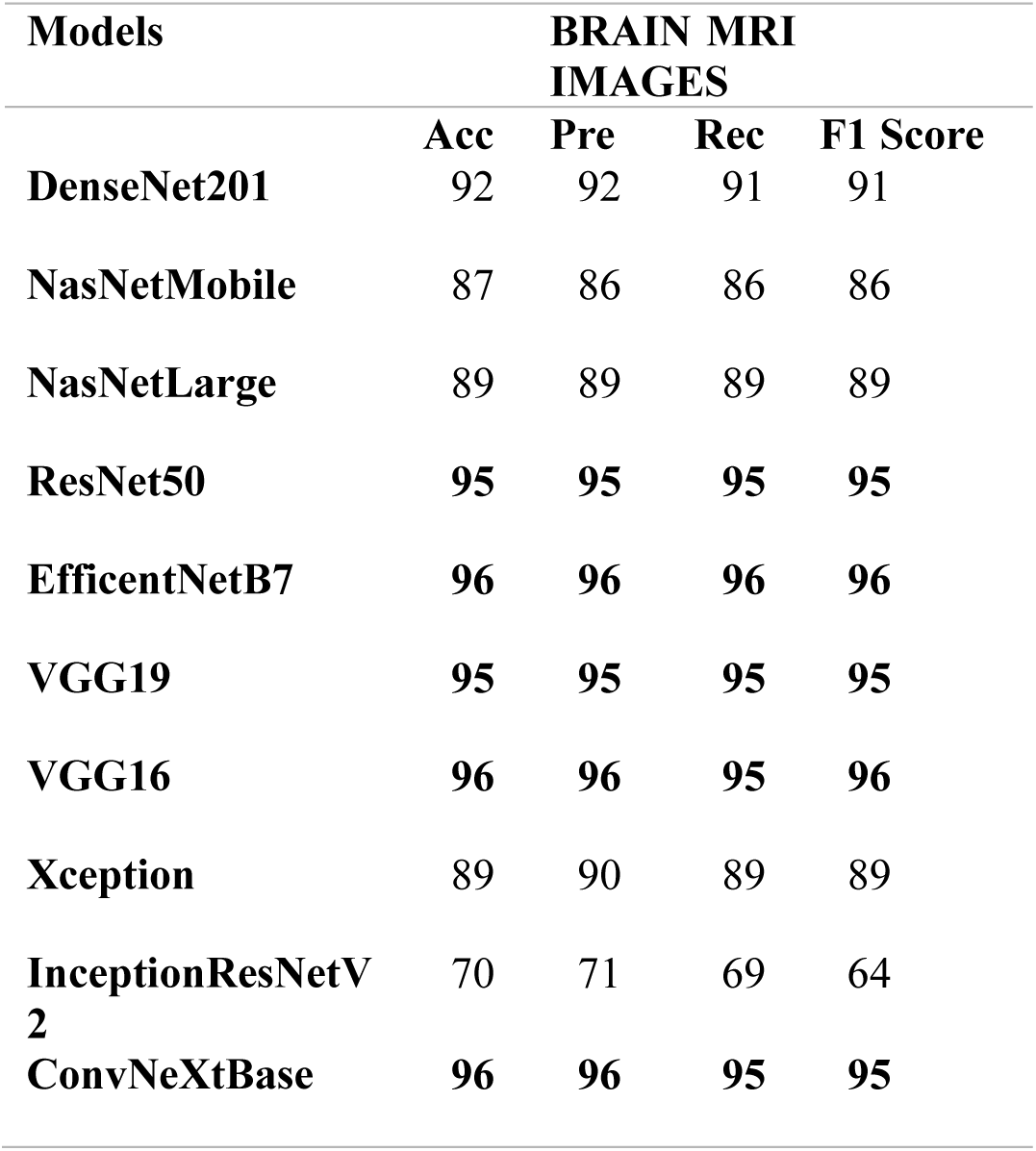
Classification Result of State of Art Pretrained Models with MRI Image Modality.

| Models | BRAIN MRI IMAGES |  |  |  |
| --- | --- | --- | --- | --- |
|  | Acc | Pre | Rec | F1 Score |
| DenseNet201 | 92 | 92 | 91 | 91 |
| NasNetMobile | 87 | 86 | 86 | 86 |
| NasNetLarge | 89 | 89 | 89 | 89 |
| ResNet50 | 95 | 95 | 95 | 95 |
| EfficientNetB7 | 96 | 96 | 96 | 96 |
| VGG19 | 95 | 95 | 95 | 95 |
| VGG16 | 96 | 96 | 95 | 96 |
| Xception | 89 | 90 | 89 | 89 |
| InceptionResNetV2 | 70 | 71 | 69 | 64 |
| ConvNeXtBase | 96 | 96 | 95 | 95 |

### 2.4 Evaluation Metrics

The metrics used for the evaluation of the models in this study are presented as follows: *TP, TN, FP, FN is True Positive, True Negative, False Positive, False Negative respectively*.

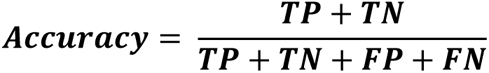

Accuracy measures how correct the model is in predicting and classifying both the positive and negative instance in the dataset.

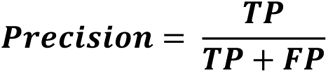

Precision specifically measures how accurate the model’s capability to precisely identify the true positive instances among all the positive predictions it made.

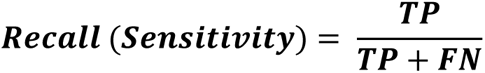

Recall, also known as sensitivity, measures how many positive instances the model accurately predicts in the dataset.

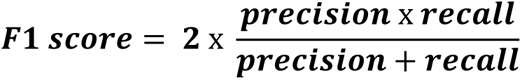

F1 Score is the harmonic mean and balance of precision and recall metrics. It takes into account both false positives (precision) and false negatives (recall), providing a balanced assessment of the model’s performance.

### 3.0 Results

This section presents the evaluation of selected models across diverse modalities, employing consistent training hyperparameters for reproducibility (outlined in Table 3). Accuracy, Precision, Recall, and F1-Score were computed for each class and averaged across the dataset, offering a comprehensive performance overview. Detailed in the tables below are the experiment results, comparing the reproducibility and generalizability of state-of-the-art CNN models. Additionally, our performance is discussed in comparison with benchmarked pretrained models done by other researchers, across Brain MRI, Kidney CT, and CXR Imaging Modalities.

**Figure 11:**
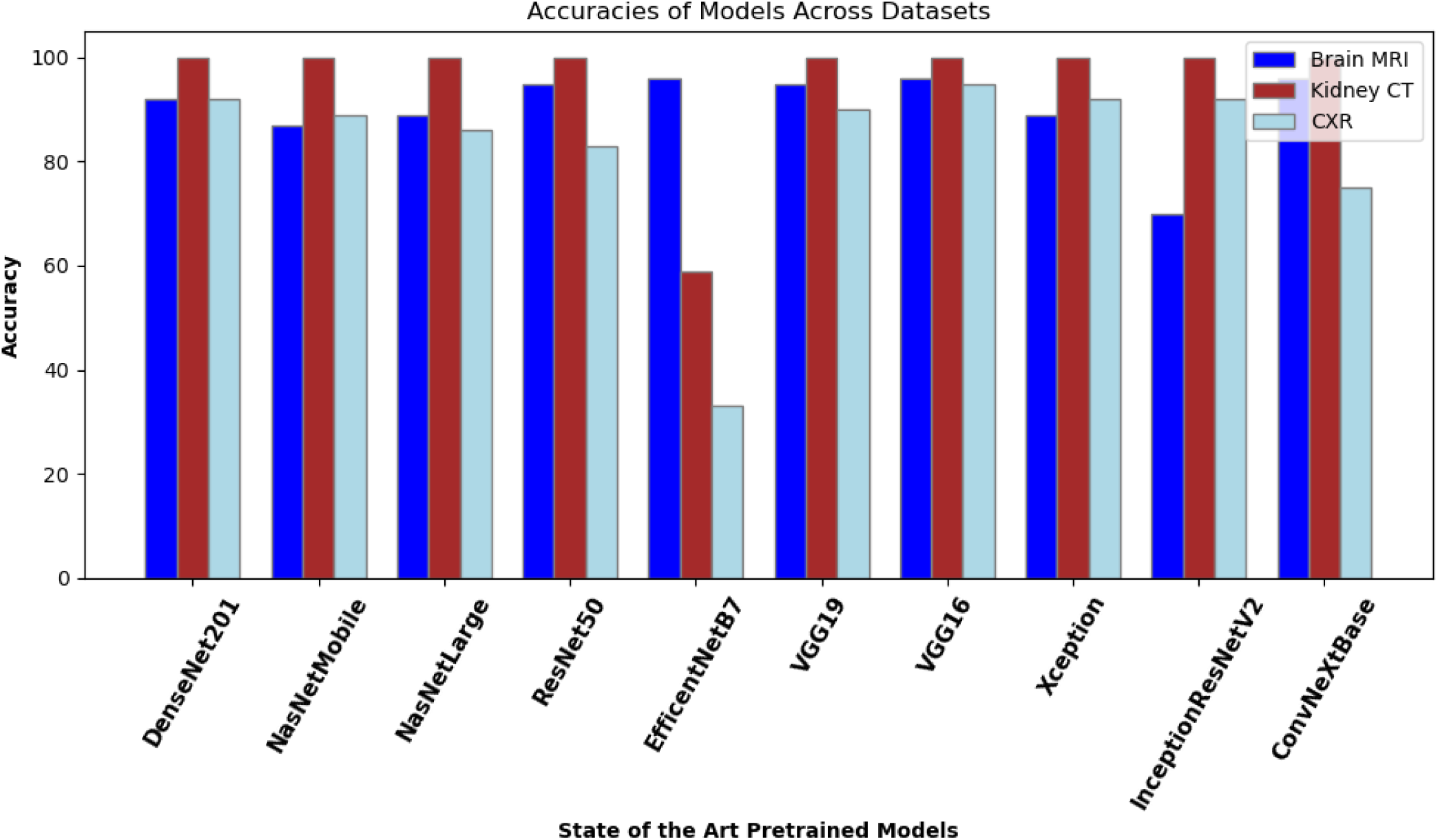
Evaluation Performance of State of Art Pretrained Models on the 3 Imaging Modalities Based Accuracy Metrics.

**Figure 12:**
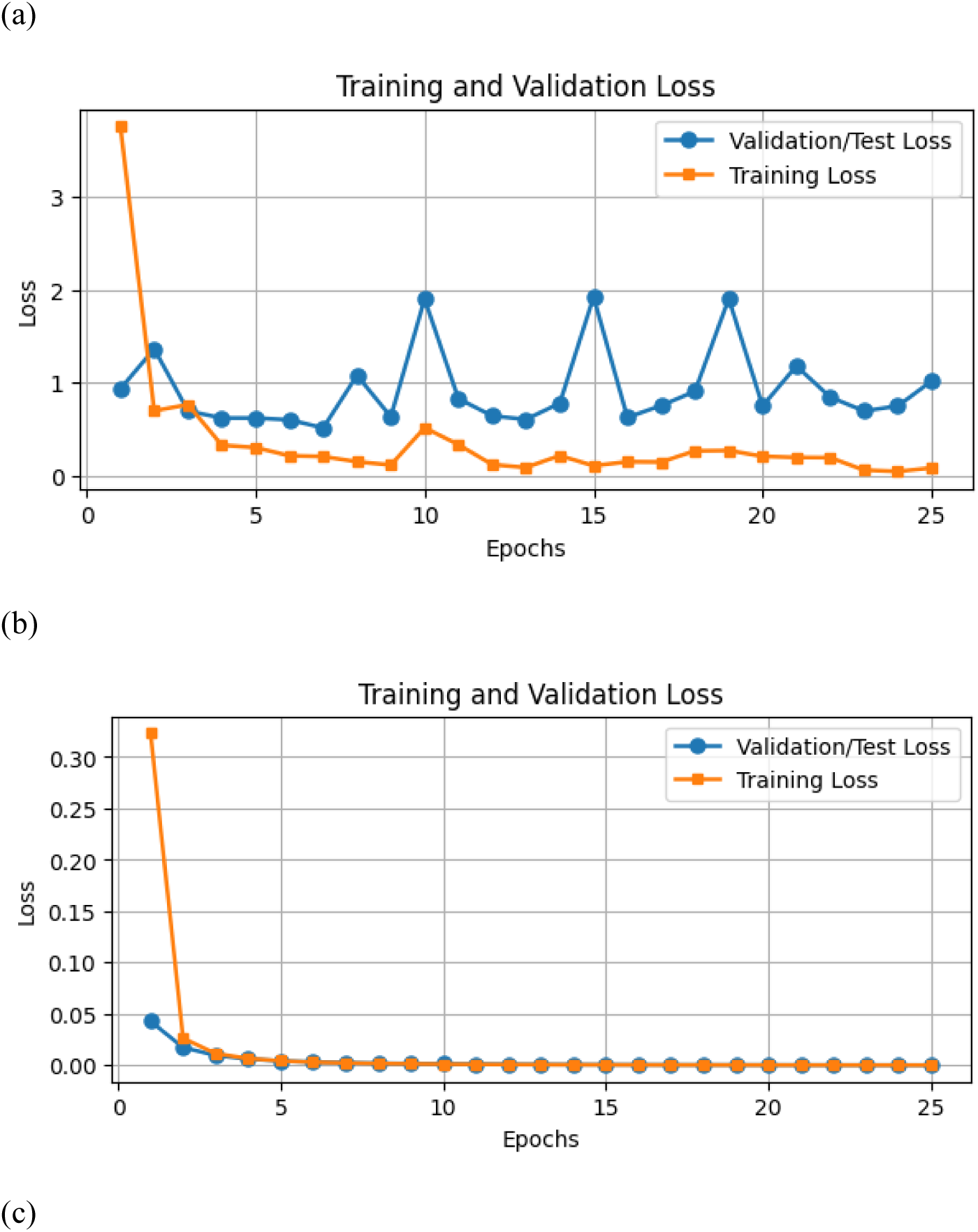
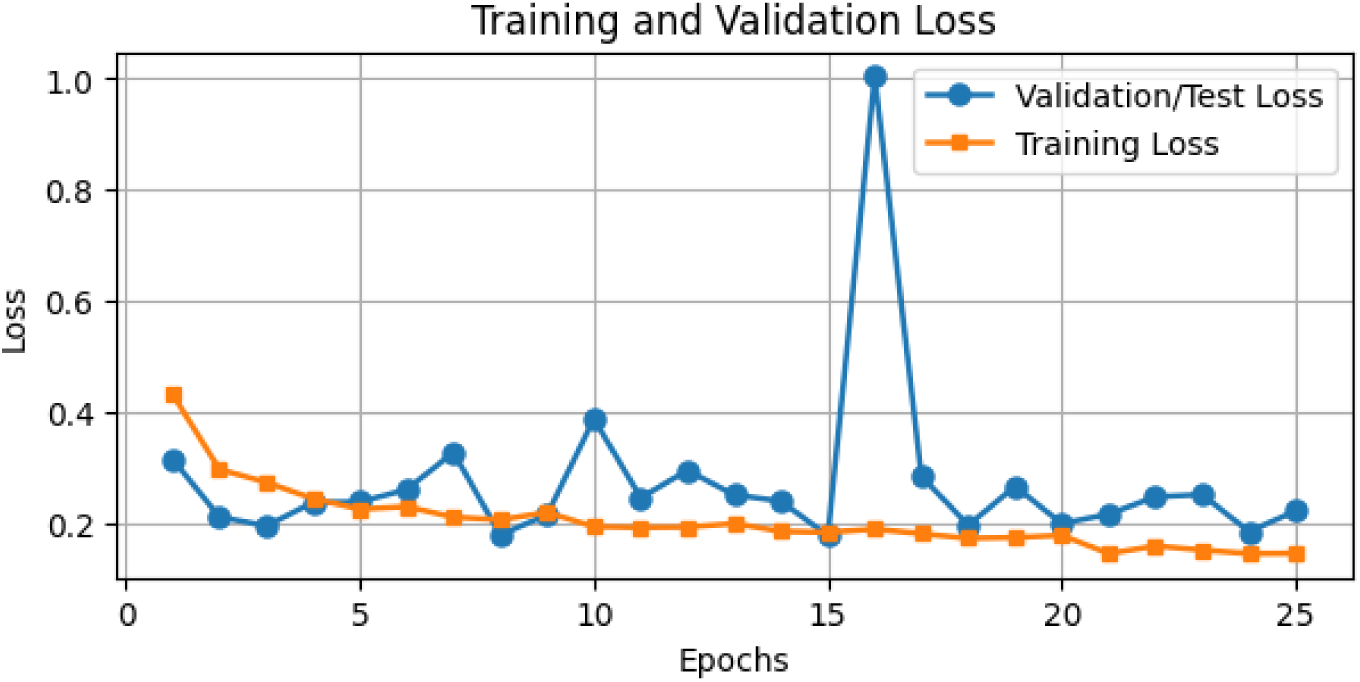
a, b, and c represent the Loss curve of VGG16 in order of MRI, CT scan and CXR modalities.

**Figure 13:**
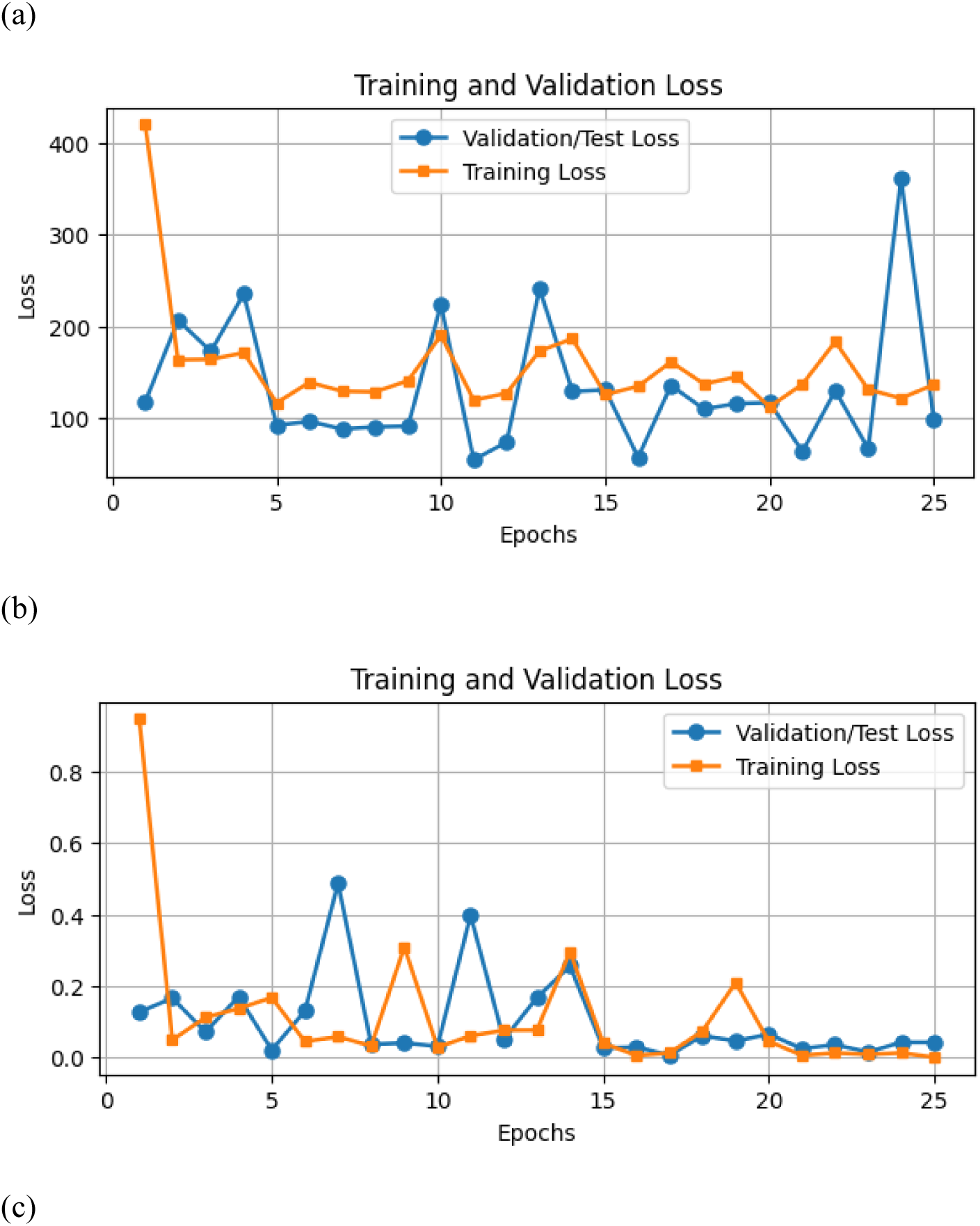
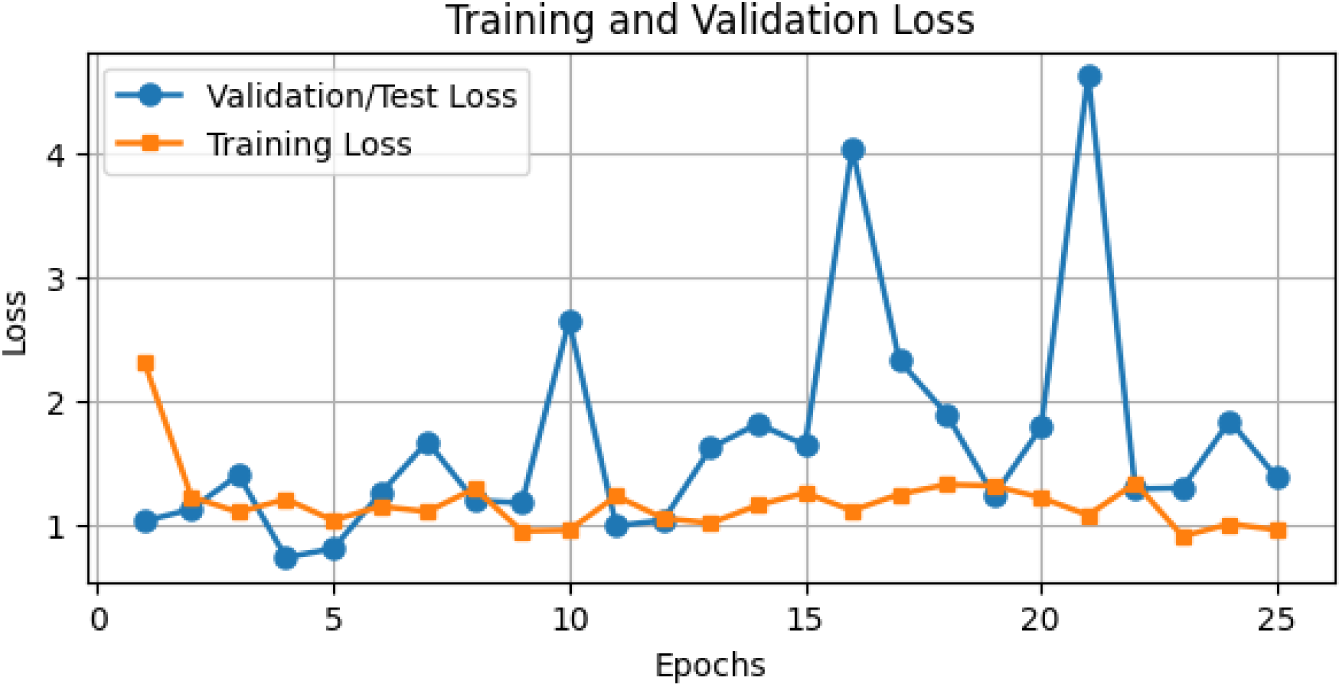
a, b, and c represent the Loss curve of InceptionResNetV2 in order of MRI, CT scan and CXR modalities.

In this experiment, ten state-of-the-art CNN models were utilized for classification across three distinct imaging modalities: brain MRI (categorizing glioma, meningioma, no tumor, and pituitary), kidney CT (identifying cyst, stone, normal, and tumor), and chest X-ray (distinguishing covid, pneumonia, and normal conditions). The results outlined in Tables 4, 5, and 6 present the top-performing models, highlighting their respective modalities. Among these models, VGG16 consistently demonstrated notable performance in identifying the various diseases in the brain, kidney and chest as provided in the dataset across the modalities, achieving 96%, 100%, and 95% accuracy in MRI, CT, and CXR classifications, respectively. While EfficientNetB7 exhibited superior recall in MRI (96% compared to VGG16’s 95%), its performance notably declined for CXR image classification as well classifying the kidney diseases with accuracies of 33% and 53% respectively. ConvNeXtBase, InceptionResNetV2, NasNetLarge, Xception exhibited inconsistent performance. On the other hand, models like DenseNet201, ResNet50, and VGG19 consistently showed high performance in MRI, CT, and CXR, although not always ranking as the top-performing models. For instance, DenseNet201 achieved 92%, 100%, and 92%, ResNet50 reached 95%, 100%, and 83%, and VGG19 attained 95%, 100%, and 90% accuracy across the modalities, respectively. Please note that the loss curves obtained for most of the pretrained models evaluated have been included under the appendix section of this paper.

**Figure 14:**
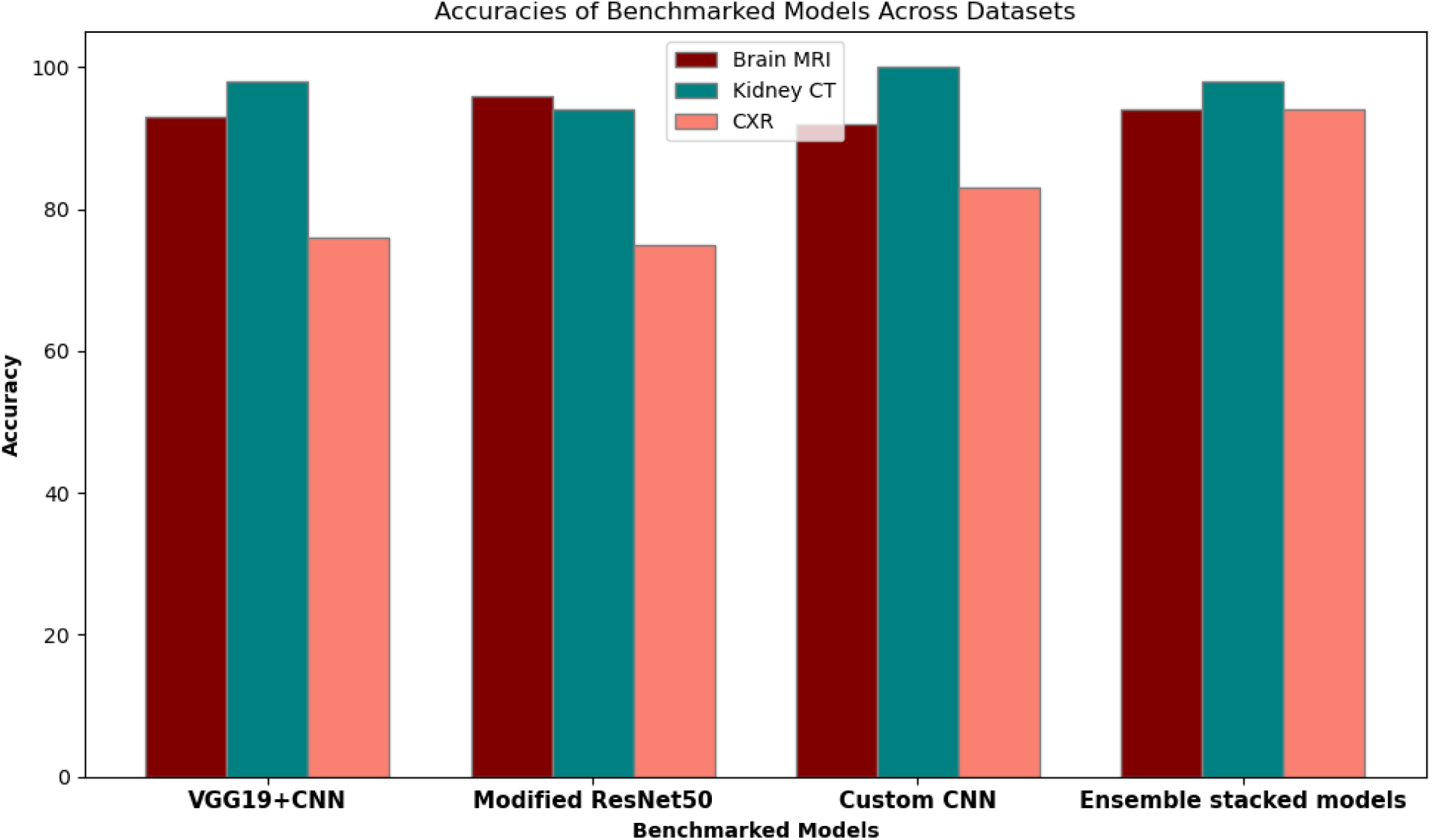
Evaluation Performance of Benchmarked Architecture on the 3 Imaging Modalities Based Accuracy Metrics.

**Table 5.** Classification Result of State of Art Pretrained Models with CT Image Modality.

| Models | KIDNEY CT IMAGES |  |  |  |
| --- | --- | --- | --- | --- |
|  | Acc | Pre | Rec | F1 Score |
| DenseNet201 | 100 | 100 | 100 | 100 |
| NasNetMobile | 100 | 100 | 100 | 100 |
| **NasNetLarge** | 100 | 100 | 100 | 100 |
| **ResNet50** | 100 | 100 | 100 | 100 |
| **EfficientNetB7** | 53 | 69 | 36 | 32 |
| **VGG19** | 100 | 100 | 100 | 100 |
| **VGG16** | 100 | 100 | 100 | 100 |
| **Xception** | 100 | 100 | 100 | 100 |
| **InceptionResNetV2** | 100 | 100 | 100 | 100 |
| **ConvNeXtBase** | 100 | 100 | 100 | 100 |

**Table 6.** Classification Result of State of Art Pretrained Models with CXR Image Modality.

| **Models** | **CXR**<br>**IMAGES** |  |  |  |
| --- | --- | --- | --- | --- |
|  | **Acc** | **Pre** | **Rec** | **F1 Score** |
| **DenseNet201** | **92** | **92** | **92** | **92** |
| **NasNetMobile** | 89 | 90 | 89 | 89 |
| **NasNetLarge** | 86 | 87 | 86 | 86 |
| **ResNet50** | 83 | 85 | 83 | 83 |
| **EfficientNetB7** | 33 | 44 | 33 | 17 |
| **VGG19** | 90 | 92 | 90 | 90 |
| **VGG16** | **95** | **95** | **95** | **95** |
| **Xception** | **92** | **92** | **92** | **92** |
| **InceptionResNetV2** | **92** | **93** | **92** | **92** |
| **ConvNeXtBase** | 75 | 82 | 75 | 75 |

The study incorporated model architectures from tables 7, 8, and 9, adapting their distinct hyperparameters to suit the dataset. The VGG19+CNN model, as reported by Alshmrani et al. (2023), exhibited promising performance with an accuracy of 98% on a CXR dataset. Upon evaluation across a broader spectrum of modalities encompassing MRI, CT, and CXR, our study observed accuracies of 93%, 100%, and 76% respectively. The modified ResNet50, initially assessed on two CXR datasets with accuracies of 99.2% and 86.1%, displayed accuracies of 96%, 94%, and 75% across the MRI, CT, and CXR modalities when compared against our study’s datasets.

**Table 7.** Classification Result of Evaluating Benchmarked with MRI Image Modality.

| Method | Models | Classes | Modality Used in article | Existing Accuracy | Evaluation Results |  |  |  |
| --- | --- | --- | --- | --- | --- | --- | --- | --- |
|  |  |  |  |  | Acc | Pre | Rec | F1 Score |
| Alshmrani et al. (2023) | VGG19+CNN | 6 | CXR dataset (1) | 98 | 93 | 94 | 93 | 93 |
| Agrawal et al. (2023) | Modified ResNet50 | 2 & 3 | CXR dataset (2) | 99.2, 86.1 | 96 | 96 | 95 | 96 |
| Nayak et al. (2020) | Custom CNN | 5 | MRI dataset (2) | 100, 97.5 | 92 | 92 | 91 | 92 |
| Jangam et al. (2022) | Ensemble stacked models | 2 | CXR dataset (2) & CT dataset (3) | 84, 93, 99, 99, 90 | 94 | 94 | 93 | 94 |

**Table 8.** Classification Result of Evaluating Benchmarked with CT Image Modality.

| Method | Models | Classes | Modality Used in article | Existing Accuracy | Evaluation Results |  |  |  |
| --- | --- | --- | --- | --- | --- | --- | --- | --- |
|  |  |  |  |  | Acc | Pre | Rec | F1 Score |
| Alshmrani et al. (2023) | VGG19+CNN | 6 | CXR dataset (1) | 98 | 100 | 100 | 100 | 100 |
| Agrawal et al. (2023) | Modified ResNet50 | 2 & 3 | CXR dataset (2) | 99.2, 86.1 | 94 | 94 | 91 | 92 |
| Nayak et al. (2020) | Custom CNN | 5 | MRI dataset (2) | 100, 97.5 | 100 | 100 | 100 | 100 |
| **Jangam et al. (2022)** | Ensemble stacked models | 2 | CXR dataset (2) & CT dataset (3) | 84, 93, 99, 99, 90 | 98 | 97 | 99 | 98 |

**Table 9.** Classification Result of Evaluating Benchmarked with CXR Image Modality.

| Method | Models | Classes | Modality Used in article | Existing Accuracy | Evaluation Results |  |  |  |
| --- | --- | --- | --- | --- | --- | --- | --- | --- |
|  |  |  |  |  | Acc | Pre | Rec | F1 Score |
| **Alshmrani et al. (2023)** | VGG19+CNN | 6 | CXR dataset (1) | 98 | 76 | 84 | 76 | 75 |
| **Agrawal et al. (2023)** | Modified ResNet50 | 2 & 3 | CXR dataset (2) | 99.2, 86.1 | 75 | 75 | 75 | 75 |
| **Nayak et al. (2020)** | Custom CNN | 5 | MRI dataset (2) | 100, 97.5 | 83 | 85 | 83 | 82 |
| **Jangam et al. (2022)** | Ensemble stacked models | 2 | CXR dataset (2) & CT dataset (3) | 84, 93, 99, 99, 90 | 94 | 94 | 94 | 94 |

Two MRI datasets were utilized to evaluate the custom CNN, achieving accuracies of 100% and 97.5%. The performance evaluation in our study showcased accuracies of 92%, 100%, and 83% across the MRI, CT, and CXR modalities. The ensemble methods, scrutinized across five datasets—two belonging to CXR and three to CT image modalities—maintained consistent performance with accuracies ranging from 84% to 99% for CXR datasets and from 90% to 99% for CT datasets. In contrast to our study, the ensemble method exhibited analogous results of 94%, 98%, and 94% across the MRI, CT, and CXR modalities. The ensemble method reliably maintained high accuracy across diverse image modalities. This approach utilized a fusion of multiple models, leveraging their individual strengths to collectively achieve a sustained, exceptional level of accuracy of 94%, 98%, and 94% across the diverse spectrum of medical imaging modalities under examination in this study.

## 4.0 Discussion

The outcome of this study has provided insight into the adaptability of pre-trained models across MRI scans, CT scans, and X-rays, spotlighting VGG16 as a standout performer across Brain MRI, Kidney CT, and CXR datasets, boasting impressive accuracies of 96%, 100%, and 95% respectively. Insights from implementing the ensemble method from existing literature reveals the ensemble method exhibited a sustained exceptional accuracy of 94%, 98%, and 94% for Brain MRI, Kidney CT, and CXR images respectively, showcasing its potential in achieving adaptability across diverse modalities. In contrast to the adaptability of Vgg16, the EfficientNetB7 model demonstrated high performance specifically in Brain MRI classification with 96% accuracy, its performance notably dipped when evaluated on the Kidney CT and CXR images with accuracy of 53% and 33% respectively. The result of efficientNetB7 based on the findings of similar studies by Abirami (2023), where EfficientNetB7 model performed with 98.4% accuracy in classifying four classes of brain MRI diagnosis might suggest the potential specialization for MRI classification specifically. These results build on existing need of ensuring consistent performance across diverse datasets and imaging modalities, crucial for dependable clinical decision-making and mitigating bias present in pretrained models which this study uncovers. While the future of AI in medical image interpretation holds immense potential, certain limitations merit attention. The rapid evolution of AI models may challenge the enduring relevance of findings, emphasizing the need for ongoing assessment and adaptation to emerging methodologies. Additionally, the study’s focused approach on specific modalities might inadvertently overlook emerging ones, potentially limiting the broader applicability of findings to a more extensive array of pretrained models not encompassed in the analysis. This study emphasizes the importance of validating pretrained models across various modalities to ensure their robustness and suitability for medical image interpretation within the healthcare sector. These findings serve as a crucial guideline for selecting appropriate models, significantly impacting their successful application in medical imaging tasks.

## 5.0 Conclusion

The evolution of pretrained models in medical image classification has significantly impacted radiology, offering a reliable means for interpretation. By analyzing these pretrained models for their efficacy across three key imaging modalities: MRI scans, CT scans, and X-rays, focusing on their adaptability, it can be concluded that pretrained models are robust in reproducing high performance across diverse datasets and imaging modalities. This study underscores the potential widespread application of the VGG16 model in interpreting various medical images for its reproducibility and generalizability, while also emphasizing the specialized use of certain pretrained models for specific imaging task like the EfficientNetB7 which specifically excelled in MRI classification. Additionally, the study highlights that although other high-performing models might not be optimal initially, fine-tuning could yield more favourable outcomes as seen in the benchmarked architectures implemented on. To better understand the implication of these results, future studies should consider the validation of the VGG16 model across a wider range of medical imaging modalities than those explored in this study. Further investigation into optimizing the EfficientNetB7 model for MRI tasks is also recommended. Moreover, validating pretrained models in real-world clinical settings remains pivotal to confirm their robustness and practical applicability. This research offers valuable insights into the potential of certain pretrained models, emphasizing their suitability for distinct imaging tasks, and also leveraging these models effectively in medical imaging, contributing to the ongoing quest for more accurate, adaptable, and widely applicable AI-enabled interpretations in healthcare.

## Data Availability

All relevant data are within the manuscript and its Supporting Information files.

https://www.kaggle.com/datasets/amanullahasraf/covid19-pneumonia-normal-chest-xray-pa-dataset

https://www.kaggle.com/datasets/nazmul0087/ct-kidney-dataset-normal-cyst-tumor-and-stone

https://www.kaggle.com/datasets/navoneel/brain-mri-images-for-brain-tumor-detection

## 7.0 Appendix

Loss curves and classification report of the pretrained models in this study

**Figure 6:**
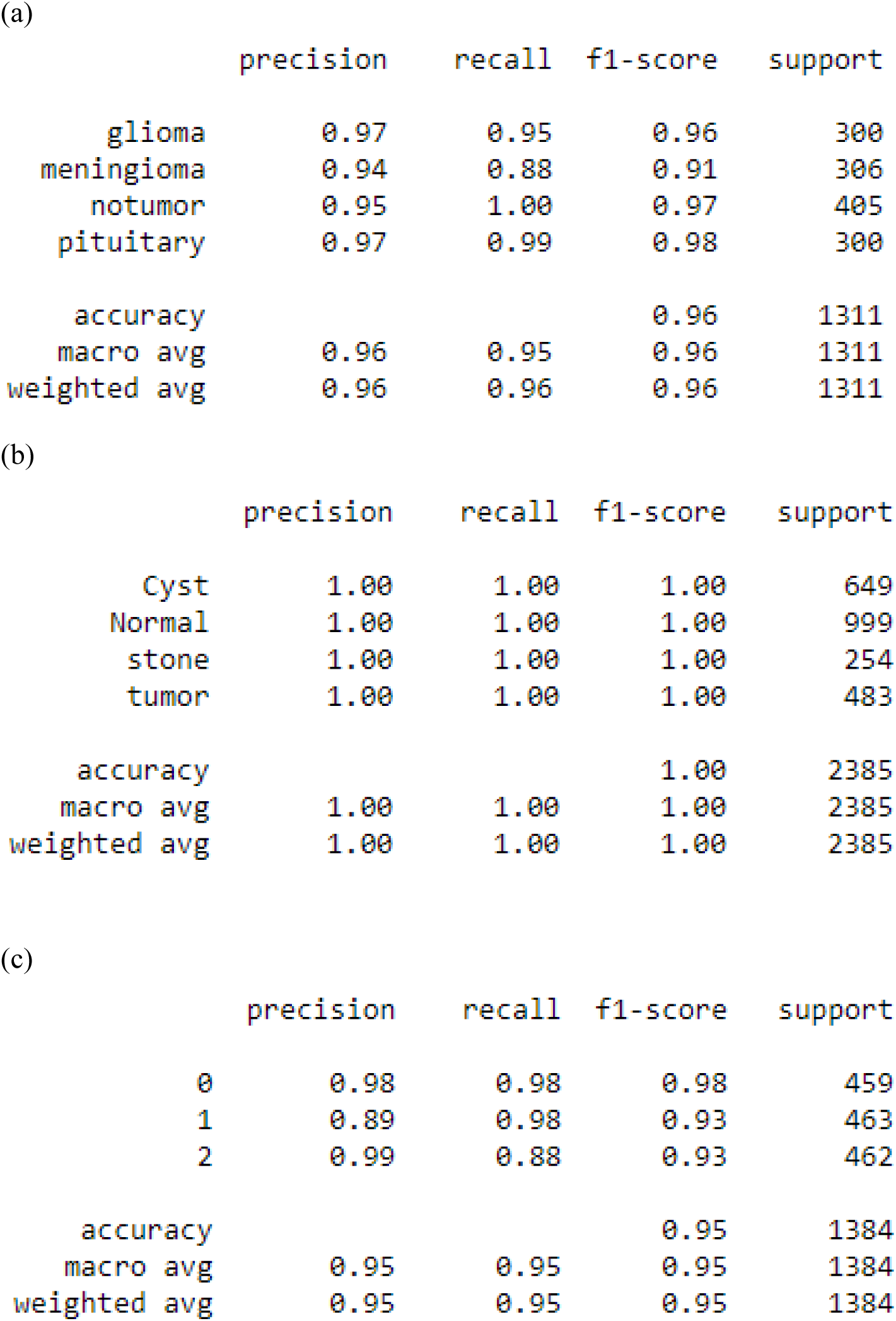
a, b, and c represent the classification report of VGG16 in order of MRI, CT scan and CXR modalities.

**Figure 11:**
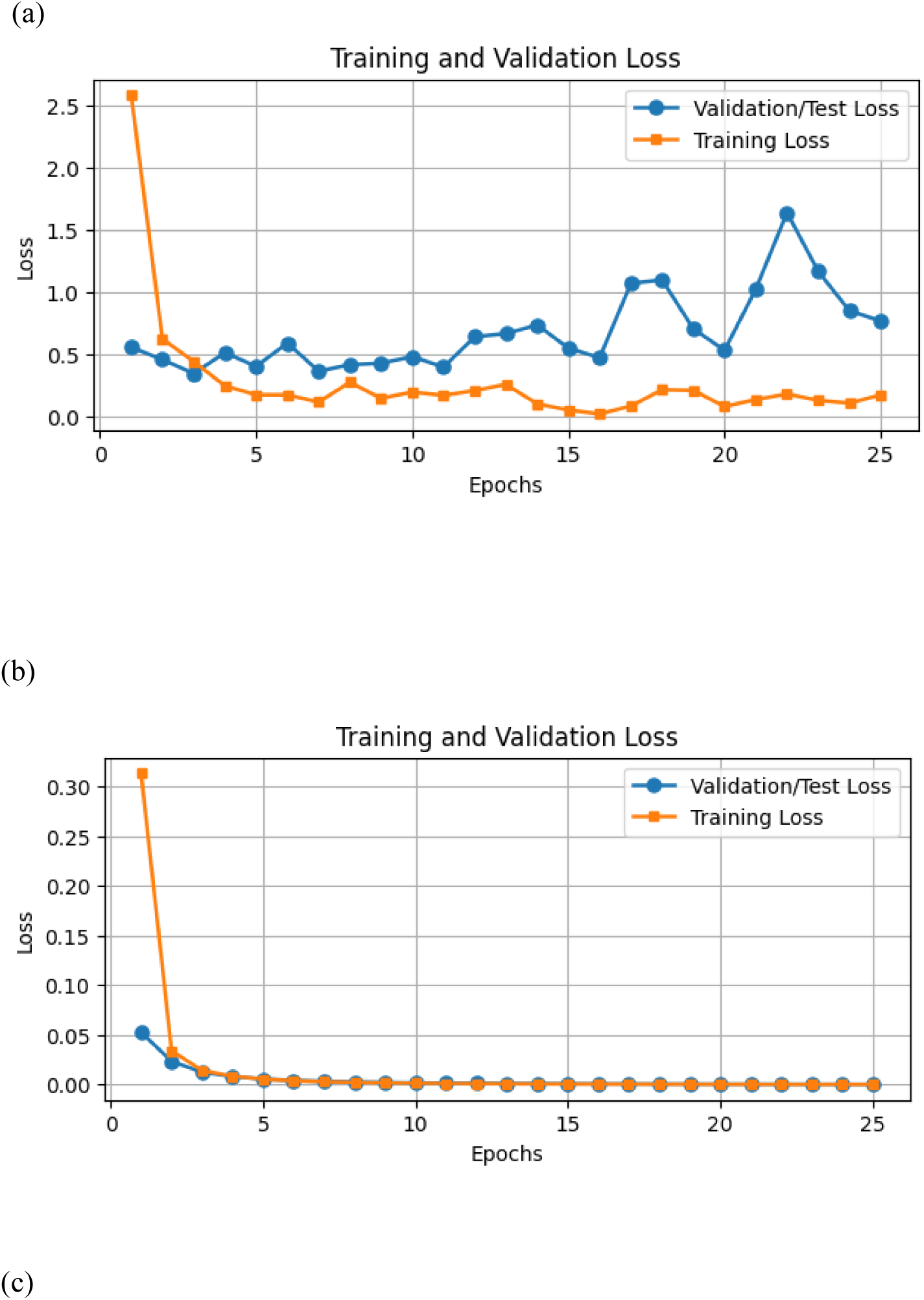
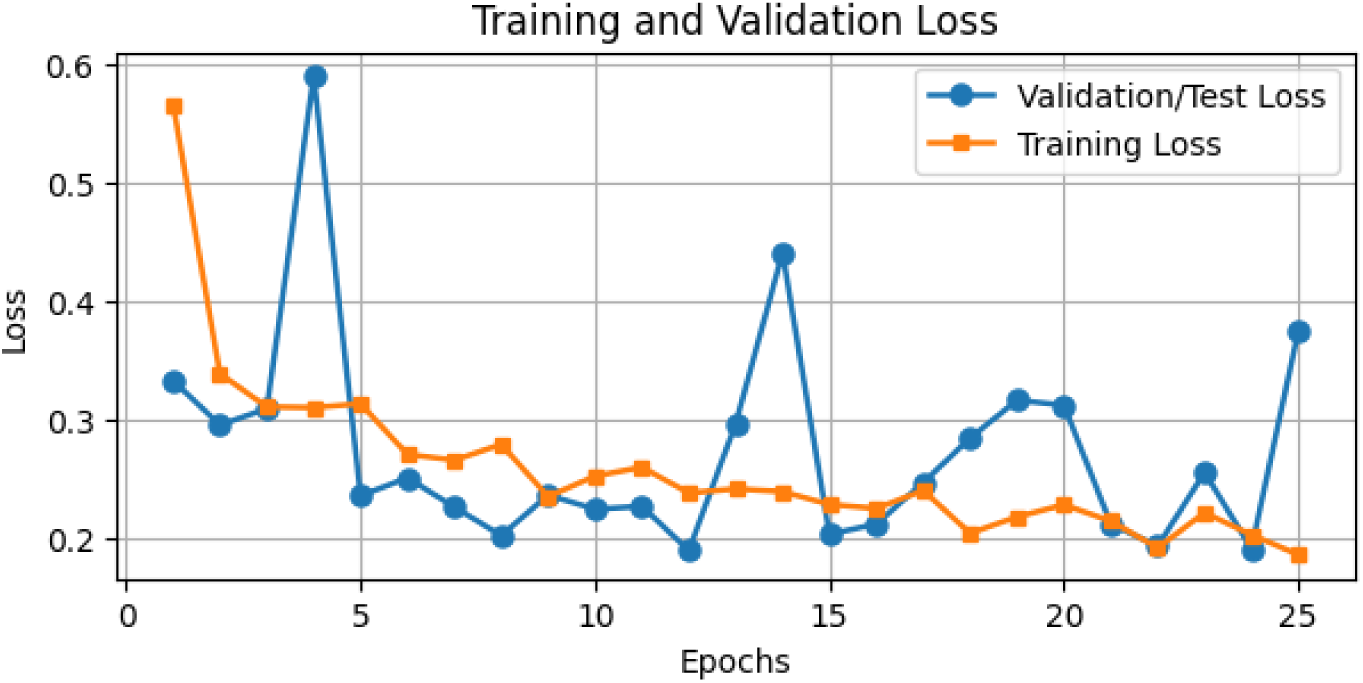
a, b, and c represent the Loss curve of VGG19 in order of MRI, CT scan and CXR modalities.

**Figure 12:**
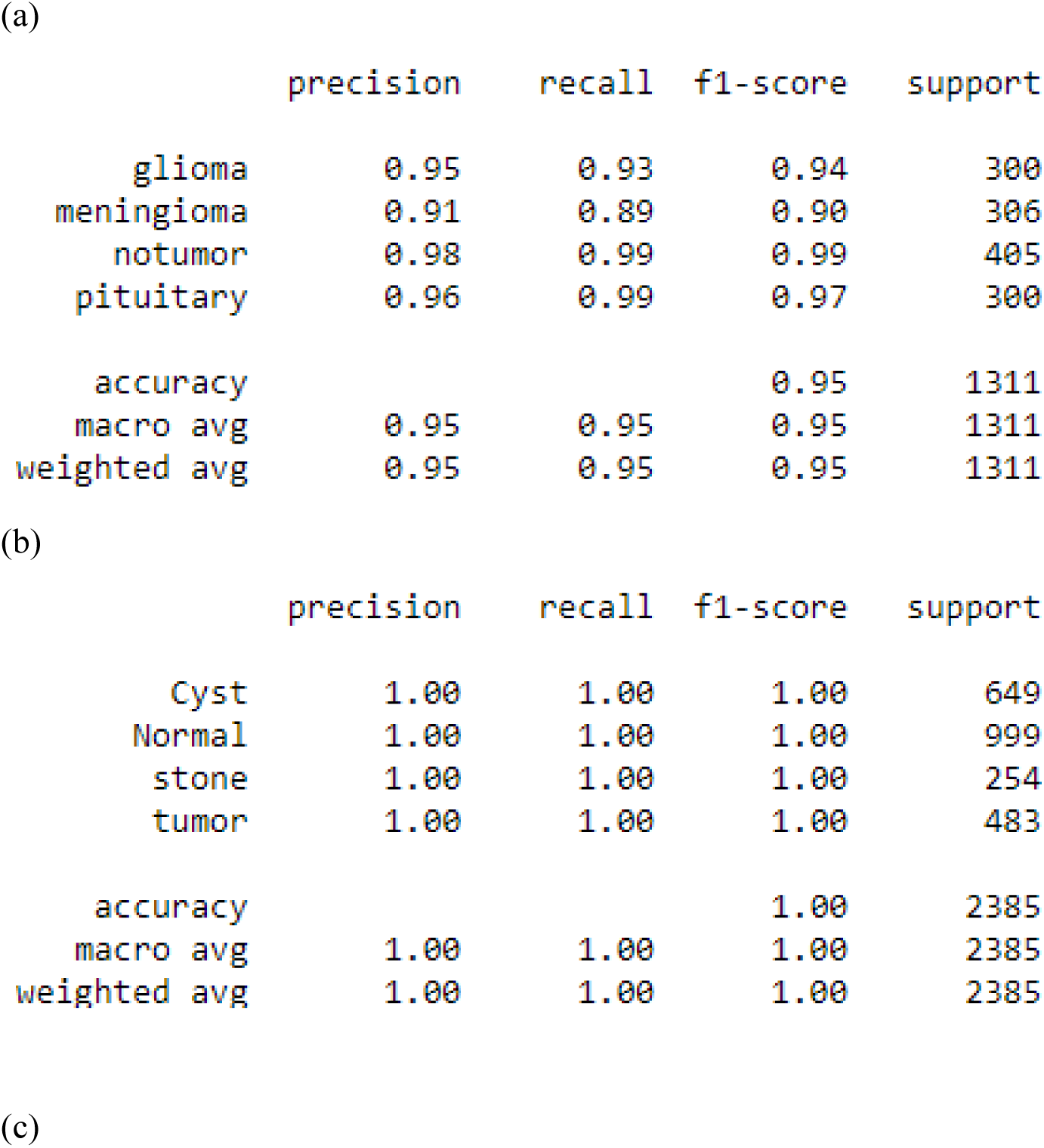
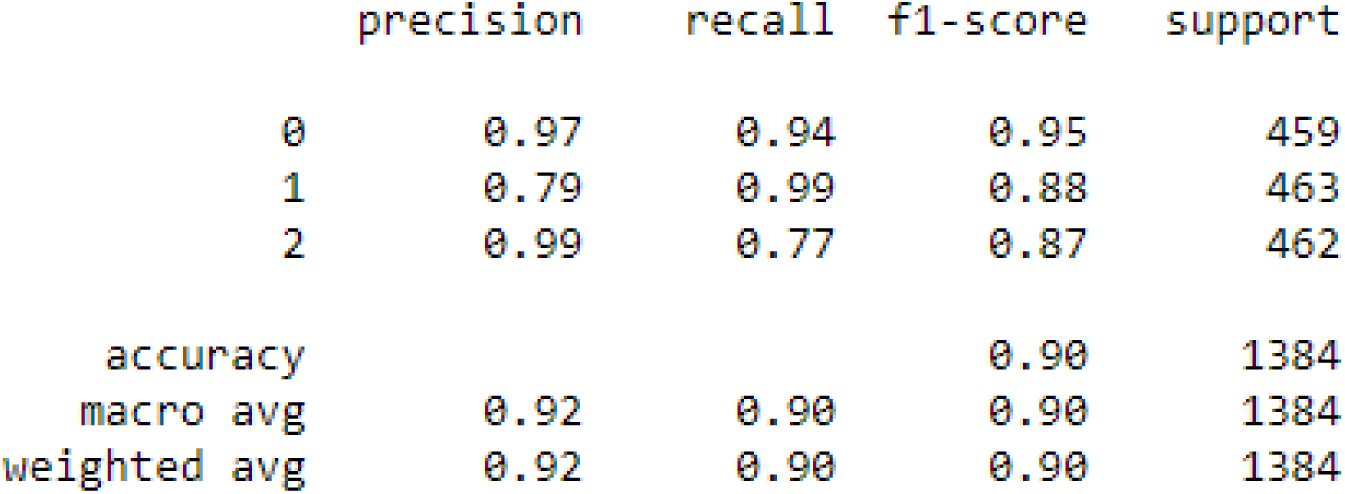
a, b, and c represent the classification report of VGG19 in order of MRI, CT scan and CXR modalities.

**Figure 13:**
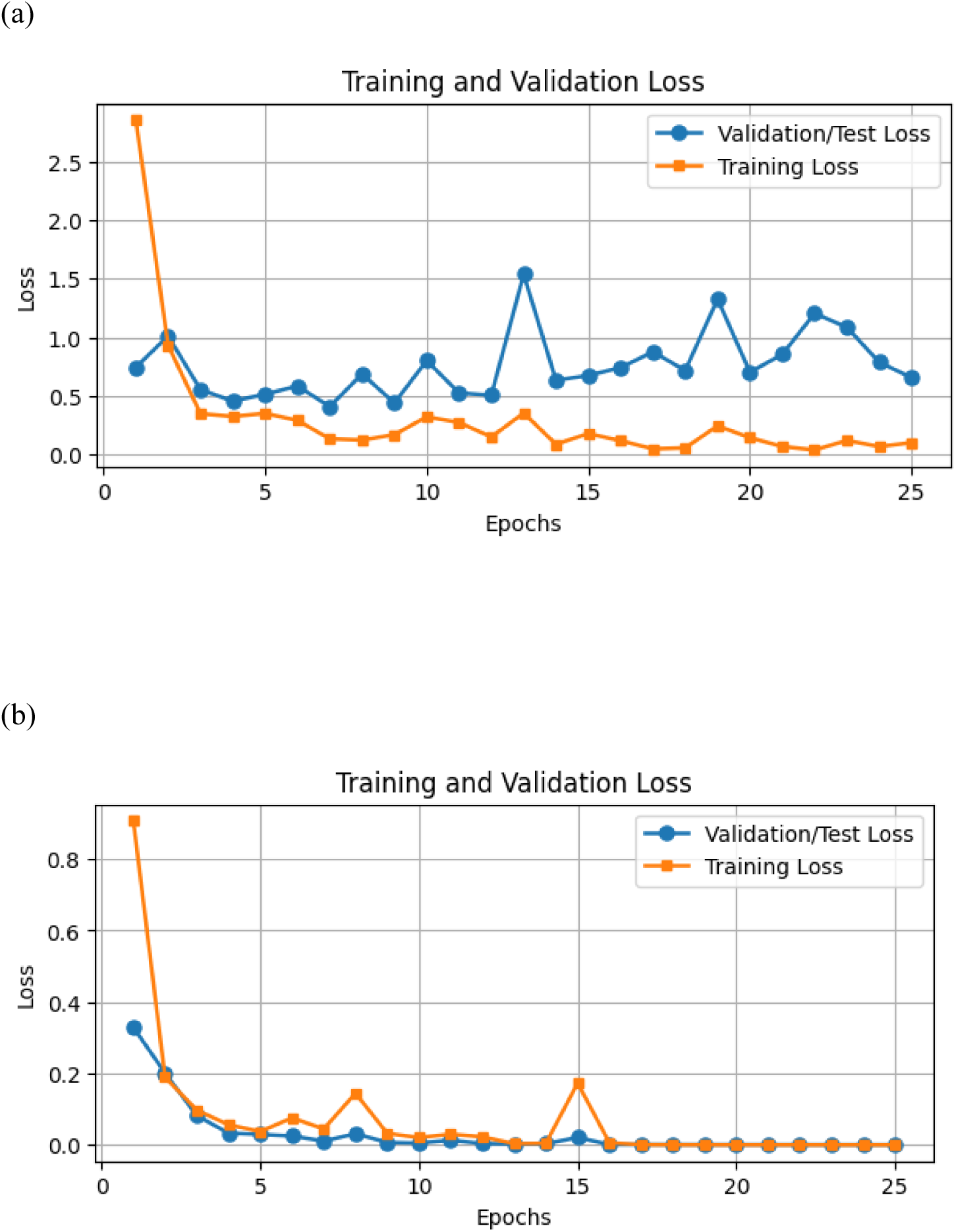
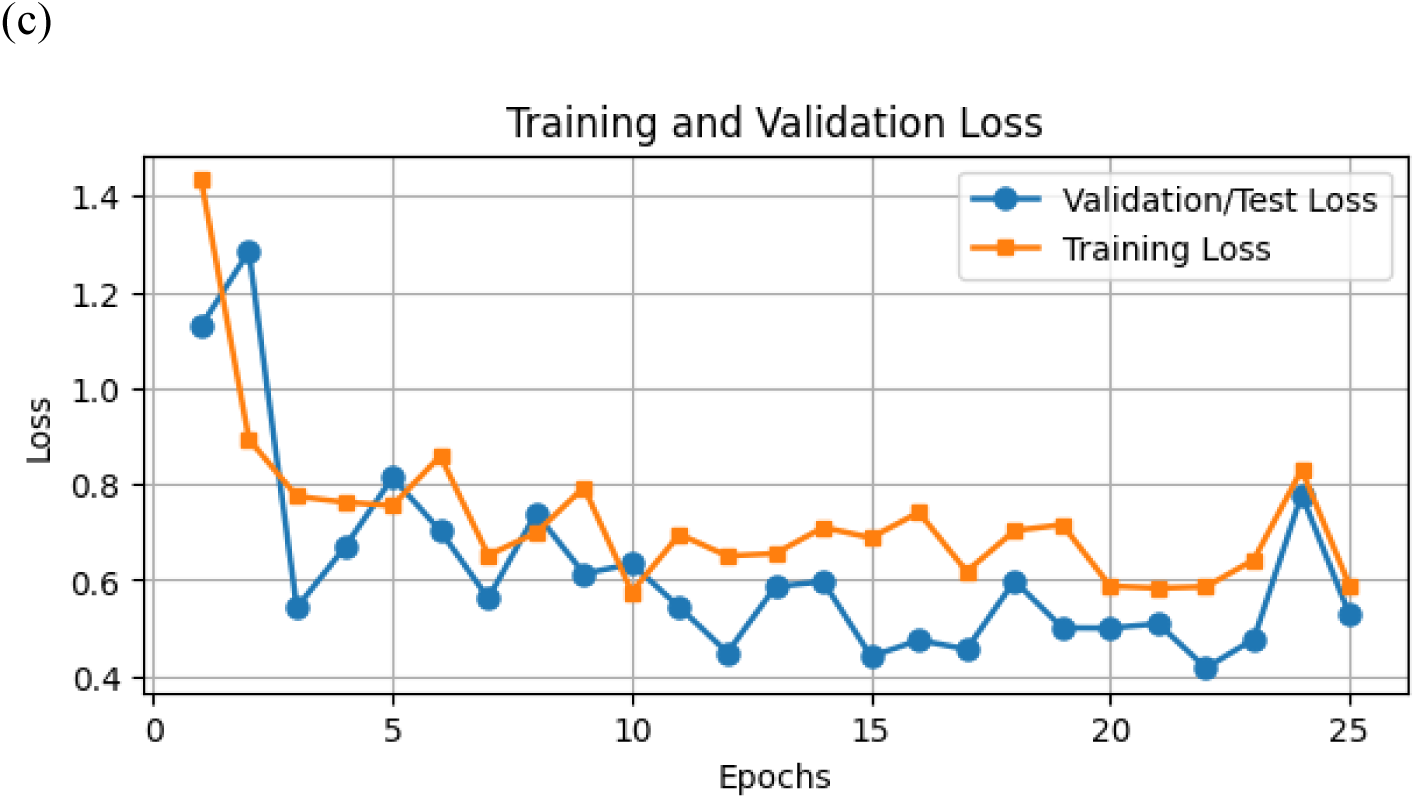
a, b, and c represent the Loss curve of ResNet50 in order of MRI, CT scan and CXR modalities.

**Figure 14:**
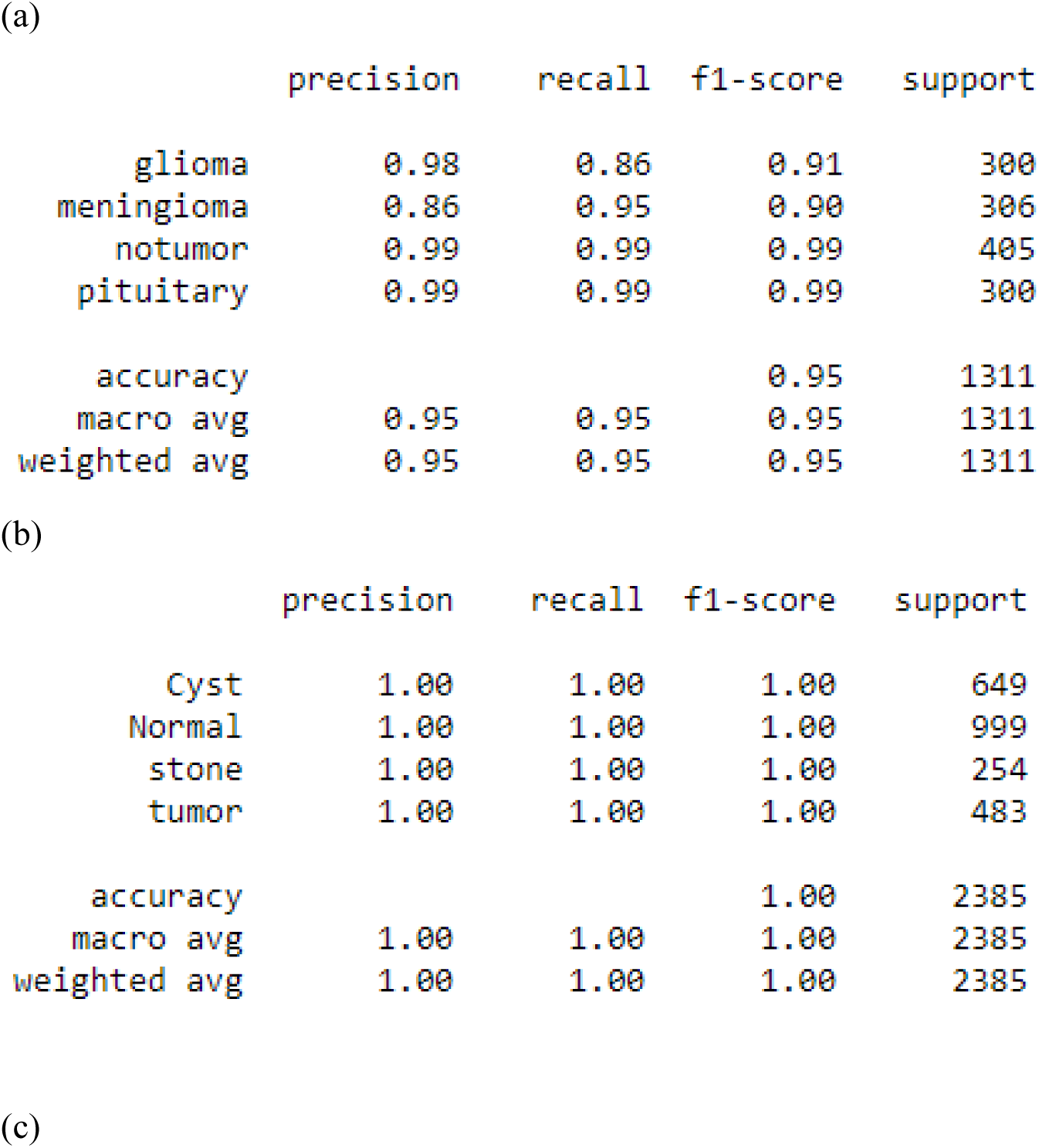
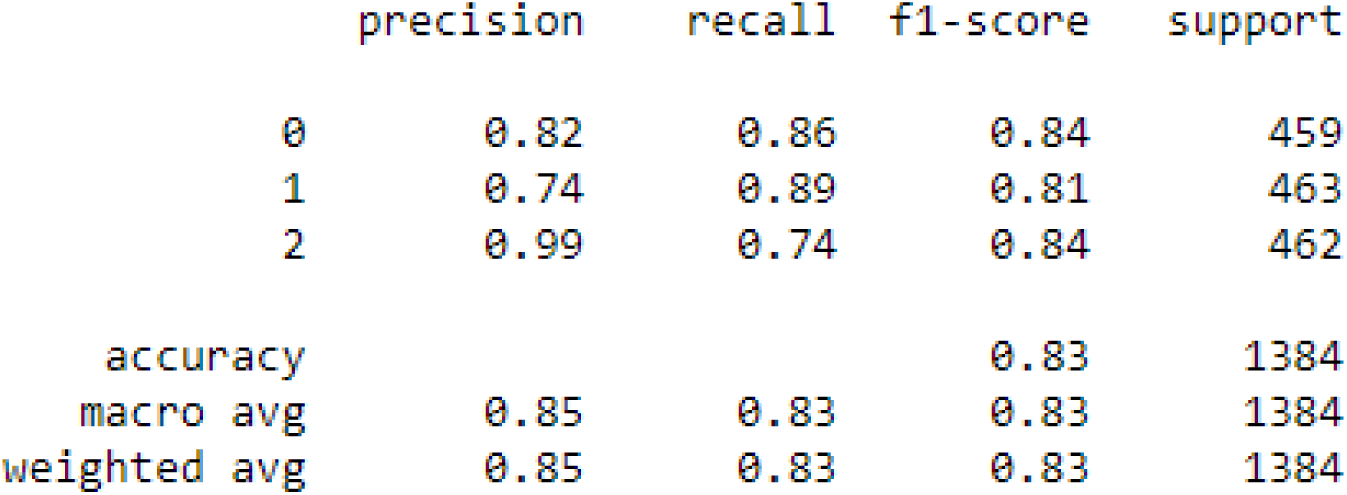
a, b, and c represent the classification report of ResNet50 in order of MRI, CT scan and CXR modalities.

**Figure 19:**
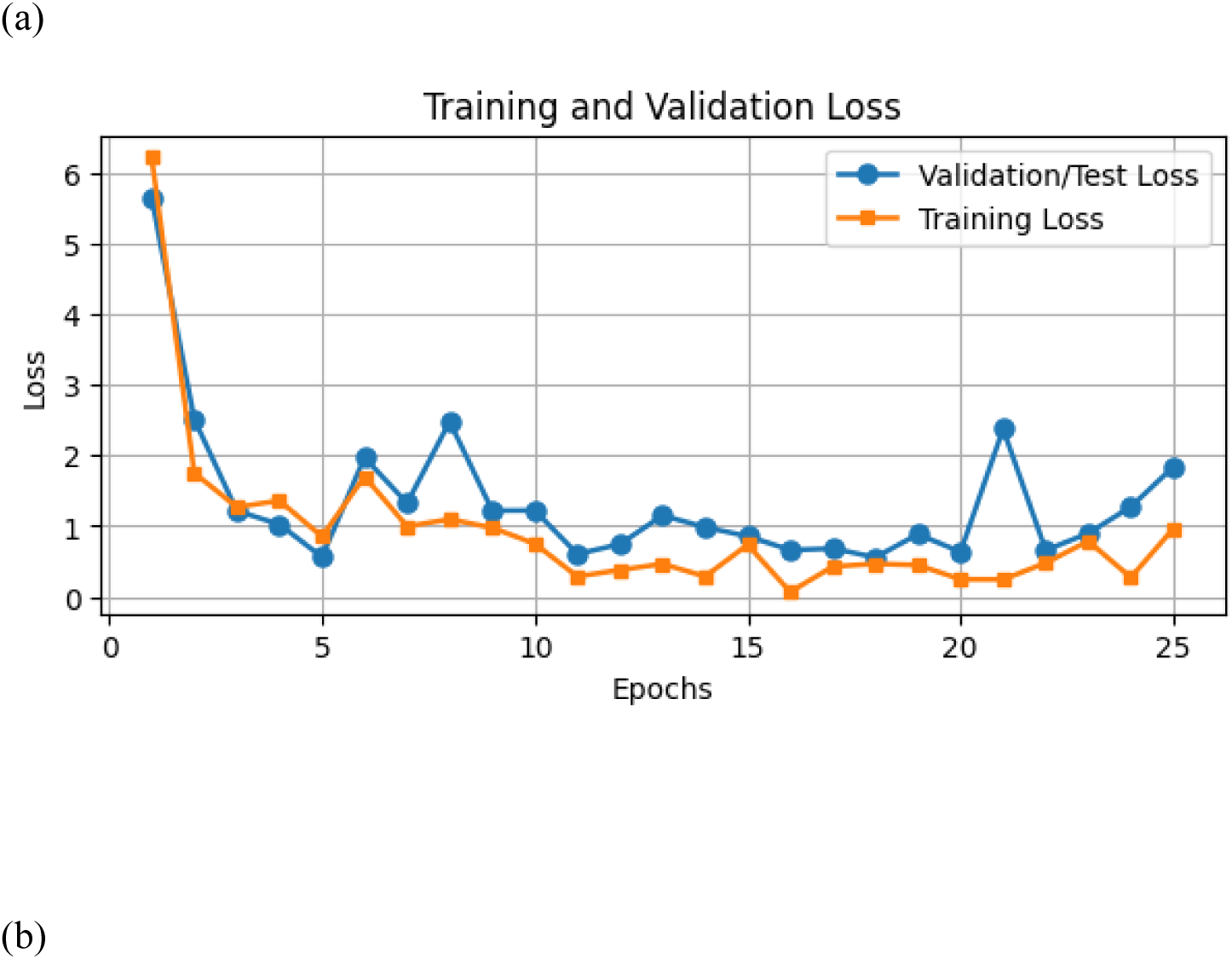
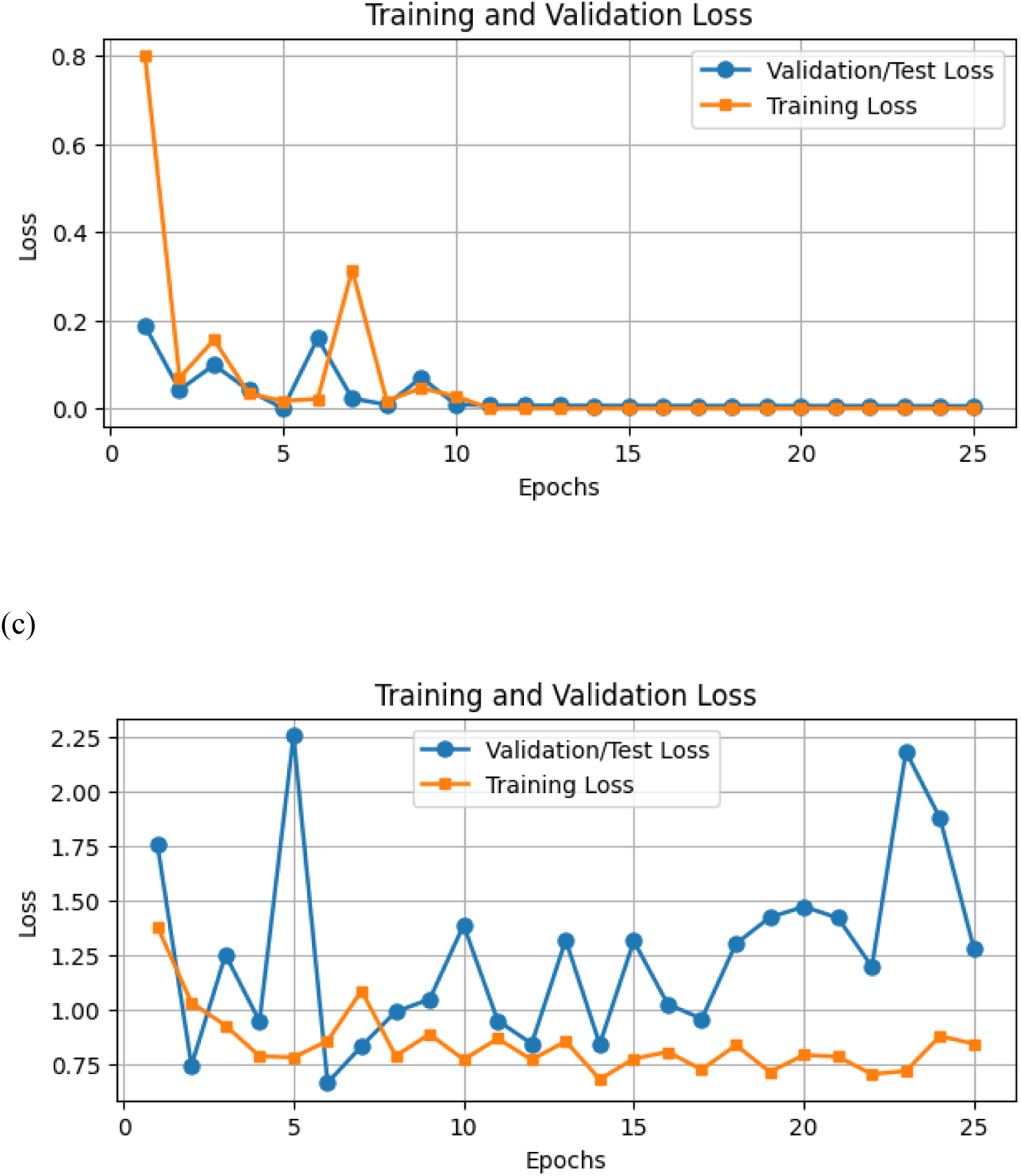
a, b, and c represent the Loss curve of DenseNet201 in order of MRI, CT scan and CXR modalities.

**Figure 20:**
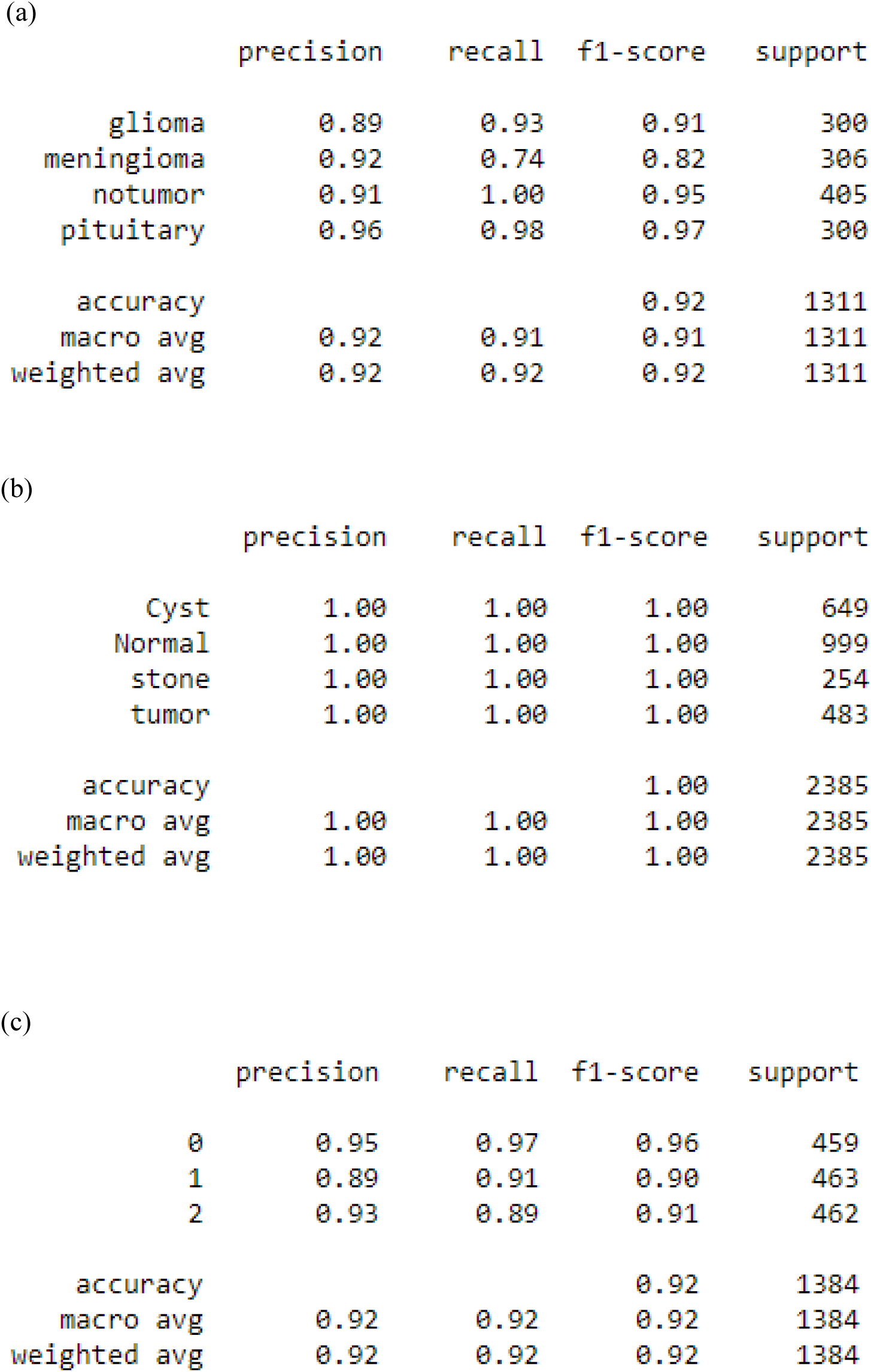
a, b, and c represent the classification report of DenseNet201 in order of MRI, CT scan and CXR modalities.

**Figure 25:**
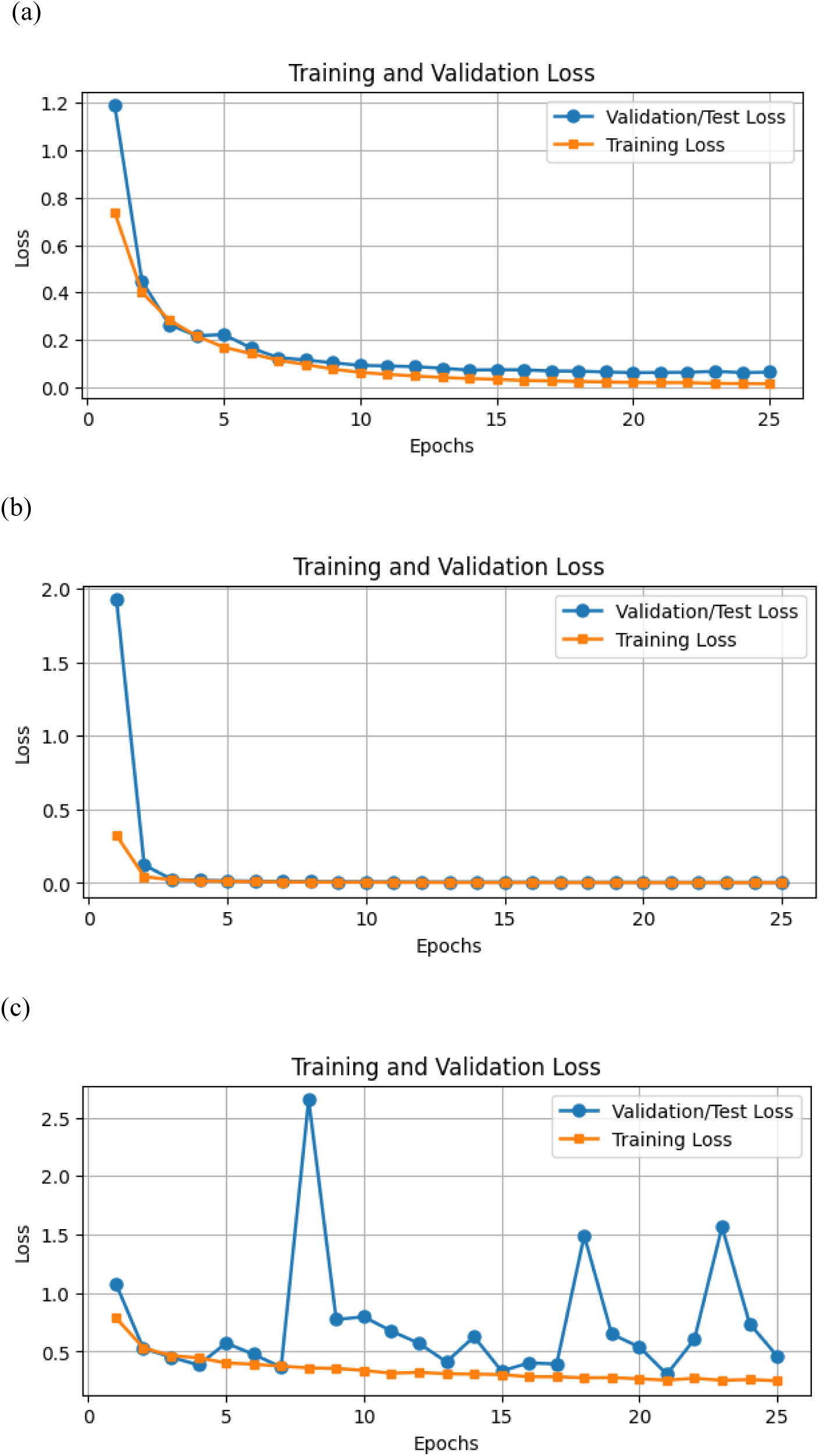
a, b, and c represent the Loss curve of CustomCNN in order of MRI, CT scan and CXR modalities.

**Figure 26:**
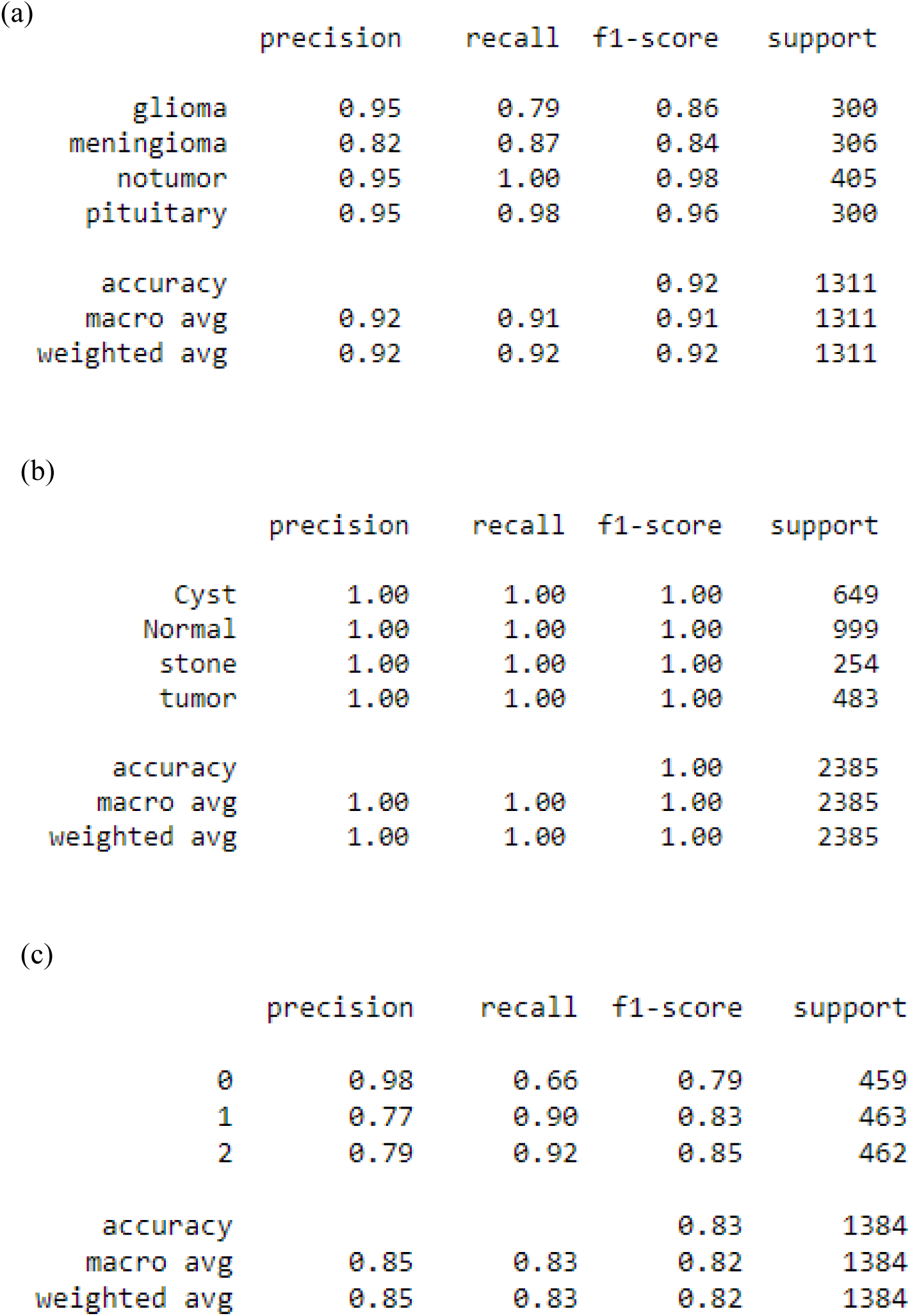
a, b, and c represent the classification report of Custom CNN in order of MRI, CT scan, and CXR modalities.

**Figure 29:**
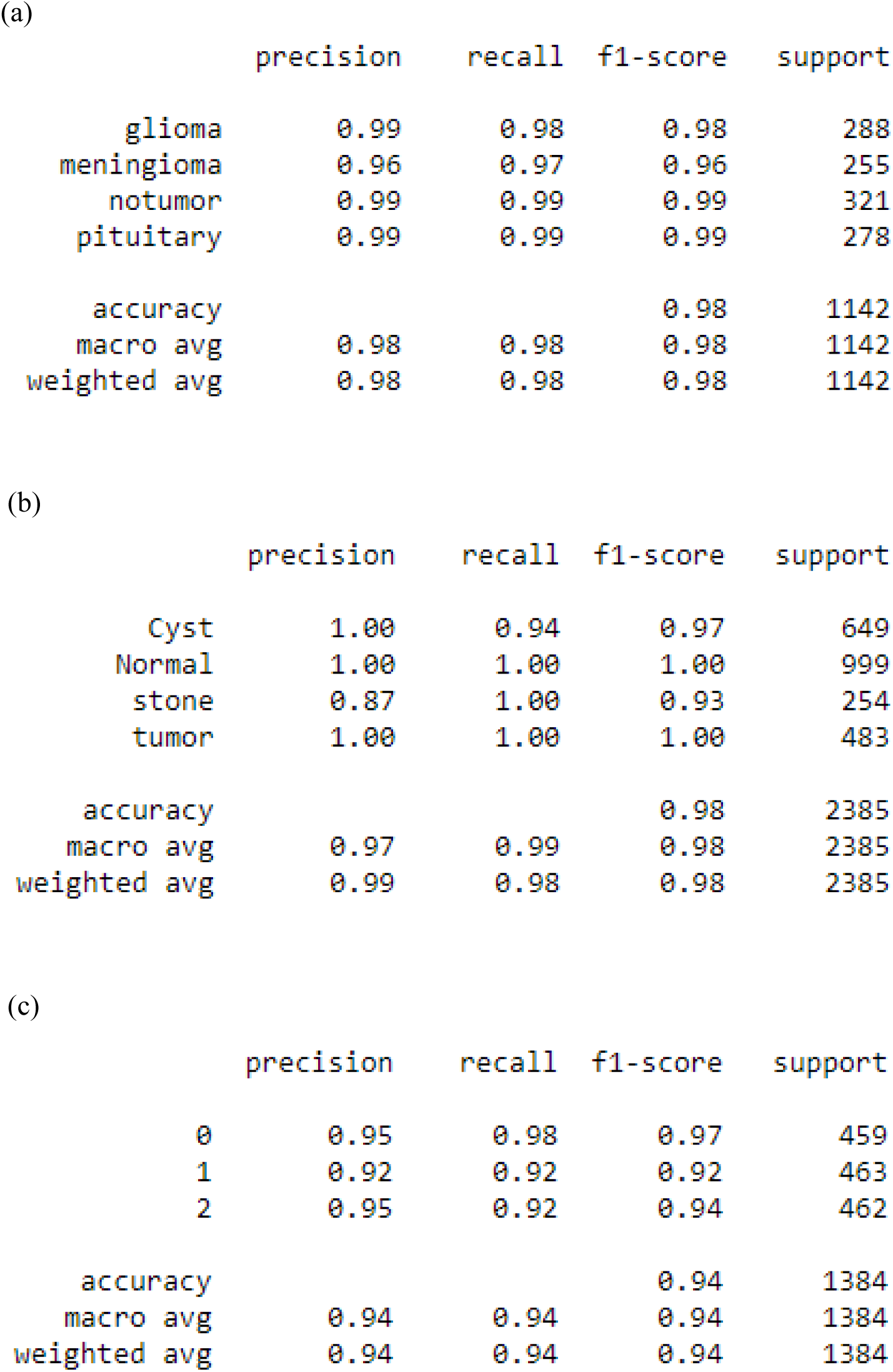
a, b, and c represent the classification report of Ensemble stacked Models in order of MRI, CT scan and CXR modalities.

## Notes

### Competing Interest Statement

The authors have declared no competing interest.

### Funding Statement

The author(s) received no specific funding for this work.

### Author Declarations

Centre of Excellence for Data Science, AI and Modelling, University of Hull, UK

